# A multi-task convolutional deep learning method for HLA allelic imputation and its application to trans-ethnic MHC fine-mapping of type 1 diabetes

**DOI:** 10.1101/2020.08.10.20170522

**Authors:** Tatsuhiko Naito, Ken Suzuki, Jun Hirata, Yoichiro Kamatani, Koichi Matsuda, Tatsushi Toda, Yukinori Okada

## Abstract

Conventional HLA imputation methods drop their performance for infrequent alleles, which is one of the factors that reduce the reliability of trans-ethnic MHC fine-mapping due to inter-ethnic heterogeneity in allele frequency spectra. We developed DEEP*HLA, a deep learning method for imputing HLA genotypes. Through validation using the Japanese and European HLA reference panels (*n* = 1,118 and 5,122), DEEP*HLA achieved the highest accuracies with significant superiority for low-frequency and rare alleles. DEEP*HLA was less dependent on distance-dependent linkage disequilibrium decay of the target alleles and might capture the complicated region-wide information. We applied DEEP*HLA to type 1 diabetes GWAS data from BioBank Japan (*n* = 62,387) and UK Biobank (*n* = 354,459), and successfully disentangled independently associated class I and II HLA variants with shared risk among diverse populations (the top signal at amino acid position 71 of HLA-DRβ1; *P* = 7.5 × 10^−120^). Our study illustrates a value of deep learning in genotype imputation and trans-ethnic MHC fine-mapping.

## Introduction

Genetic variants of the major histocompatibility complex (MHC) region at 6p21.3 contribute to the genetics of a wide range of human complex traits.^1^ Among the genes densely present in the MHC region, human leukocyte antigen (HLA) genes are considered to explain most of the genetic risk of MHC.^1^ Strategies for direct typing of HLA alleles, including sequence specific oligonucleotide (SSO) hybridization, Sanger sequencing, and next-generation sequencing (NGS), cannot be easily scaled up for large cohorts since they are labor-intensive, time-consuming, expensive, and limited in terms of allele resolution and HLA gene coverage.^2,3^ As a result, in many cases, the genotypes of HLA allele are indirectly imputed from single nucleotide variant (SNV)-level data using population-specific HLA reference panels.^3–6^ Although a high-throughput alternative is HLA type inference from whole-genome sequencing data,^7,8^ HLA imputation is still widely performed for existing single nucleotide polymorphism (SNP) genotyping data.

The MHC region harbors unusually complex sequence variations and haplotypes that are specific to individual ancestral populations; thus, the distribution and frequency of the HLA alleles are highly variable across different ethnic groups.^1,9^ This results in heterogeneity in reported HLA risk alleles of human complex diseases across diverse populations.^10^ For instance, in type 1 diabetes (T1D), the strong association between non-Asp57 in HLA-DQβ1 and T1D risk has been found in European populations^11,12^ but not in the Japanese populations, where the T1D susceptible HLA-DQβ1 alleles carry Asp57.^13^ Although the elucidation of risk alleles across ethnicities would contribute to further understanding of the genetic architecture of the MHC region associated with the pathologies of complex diseases, limited trans-ethnic MHC fine-mappings have been reported to date.^14^ One method for conducting trans-ethnic fine-mapping in the comprehensive MHC region is to newly construct a large HLA reference panel that captures the complexities of the MHC region across different populations.^15^ Another method is to integrate data of different populations that are imputed with population-specific reference panels. The latter approach appears straightforward but requires an HLA imputation method accurate enough for infrequent alleles to allow robust evaluation of HLA variants that show highly heterogenous in allele frequency across ethnicities.

Starting with a simple inference using tag SNPs,^16,17^ various methods have been developed for HLA allelic imputation. Leslie et al. first reported a probabilistic approach to classical HLA allelic imputation.^18^ HLA*IMP uses Li & Stephens haplotype model with SNP data from European populations.^19,20^ A subsequently developed software program, HLA*IMP:02, uses SNP data from multiple populations and can address genotypic heterogeneity.^21^ The current version of HLA*IMP:02 does not provide a function for users to generate an imputation model using their own reference data locally. SNP2HLA is another standard software, which uses the imputation software package Beagle to impute both HLA alleles and the amino acid polymorphisms for those classical alleles.^22^ HLA Genotype Imputation with Attribute Bagging (HIBAG)^23^ is also promising software, which employs multiple expectation-maximization-based classifiers to estimate the likelihood of HLA alleles. Whereas SNP2HLA explicitly uses reference haplotype data, for which public access is often limited, HIBAG does not require these data once the trained models are generated. These methods have achieved high imputation accuracy;^24^ however, they are less accurate for rare alleles as shown later. The complex linkage disequilibrium (LD) structures specific for the MHC region requires a more sophisticated pattern recognition algorithm beyond simple stochastic inference.

After boasting of its extremely high accuracy in image recognition, deep learning has been attracting attention in various fields. It can learn a representation of input data and extract relevant features of high complexity through deep neural networks. Many successful applications in the field of genomics have been reported.^25^ A typical application of deep learning for genomics is the prediction of the effects of non-coding and coding variants, where models encode the inputs of flanking nucleotide sequence data.^26–29^ Another application is non-linear unsupervised learning of high-dimensional quantitative data from transcriptome.^30,31^ However, successful representation learning for SNV-data in the field of population genetics is limited.^32^ Here, we developed DEEP*HLA, a multi-task convolutional deep learning method to accurately impute genotypes of HLA genes from SNV-level data. Through the application to the two HLA reference panels of different populations, DEEP*HLA achieved higher imputation accuracy than conventional methods. Notably, DEEP*HLA was advantageous especially for imputing low-frequency and rare alleles. Furthermore, DEEP*HLA showed by far the fastest total processing time, which suggests its applicability to biobank-scale data. We applied the trained models of DEEP*HLA to the large-scale T1D genome-wide association study (GWAS) data from BioBank Japan (BBJ) and UK Biobank (UKB) and conducted trans-ethnic fine-mapping in the MHC region.

## Results

### Overview of the study

An overview of our study is presented in **Fig. 1**. Our method, DEEP*HLA, is convolutional neural networks that learn from an HLA referenced panel and impute genotypes of HLA genes from pre-phased SNV data. Its framework uses a multi-task learning that can learn and impute alleles of several HLA genes which belong to the same group simultaneously (see Methods). Multi-task learning is presumed to have two advantages in this situation. First, the genotypes of some flanking HLA genes, which often show strong LD for each other, are correlated, and the shared features of individual tasks are likely to be informative. Second, the processing time is reduced by grouping tasks, especially in our latest reference panel, which comprises more than 30 HLA genes. We employed the two different HLA imputation reference panels for robust benchmarking: (i) our Japanese reference panel (*n* = 1,118)^3^ and (ii) the Type 1 Diabetes Genetics Consortium (T1DGC) reference panel (*n* = 5,122).^33^ We compared its performance with that of other HLA imputation methods by 10-fold cross-validation and an independent HLA dataset (*n* = 908).^6^ Further, we tested its imputation accuracy for multi-ethnic individuals using data from the Phase III 1000 Genomes Project (1KGv3). In the latter part, we performed MHC fine-mapping of the Japanese cohort from BBJ and British cohort from UKB by applying trained models specific for individual populations. We integrated the imputed GWAS genotypes and performed trans-ethnic HLA association analysis.

**Figure 1.**
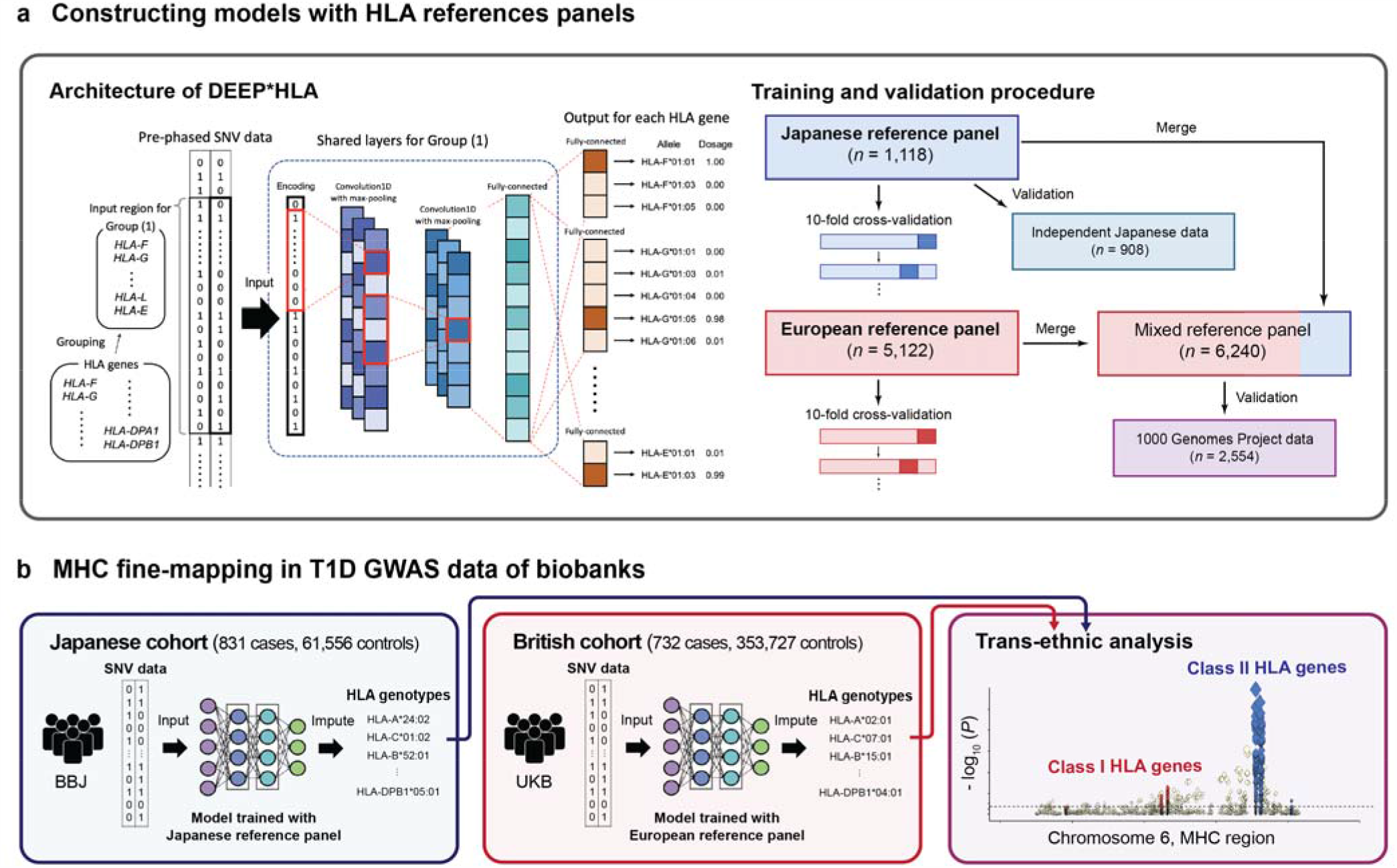
An overview of the study. (a)DEEP*HLA is a deep learning architecture that takes an input of pre-phased genotypes of SNVs and outputs the genotype dosages of HLA genes. To train a model and benchmark its performance, we used Japanese and European HLA reference panels respectively. We evaluated its accuracies in cross-validation with other methods. For the Japanese panel, we also evaluated its accuracy by applying the trained model to an independent Japanese HLA dataset. Further, we experimentally generated a mixed panel and validated its accuracy using 1KGv3 data. (**b**) We conducted trans-ethnic MHC fine-mapping in T1D GWAS data. We performed HLA imputation for the Japanese cohort from BBJ and the British cohort from UKB using models specific for individual populations. We integrated the individual results of imputed genotypes and performed trans-ethnic association analysis.

### DEEP*HLA achieved high imputation accuracy especially for low-frequency and rare alleles

First, we applied DEEP*HLA to the Japanese reference panel, a high-resolution allele catalog of NGS-based HLA typing data of the 33 classical and non-classical HLA genes along with high-density SNP data of the MHC region by genotyping with the Illumina HumanCoreExome BeadChip for 1,118 individuals of Japanese ancestry.^3^ We compared the imputation accuracy of DEEP*HLA in terms of sensitivity, positive predictive value (PPV), and *r*^*2*^ of allelic dosage, and concordance rate of best-guess genotypes (see **Methods**) with those of SNP2HLA and HIBAG in 10-fold cross-validation. DEEP*HLA achieved total sensitivity of 0.987, PPV of 0.986, *r*^*2*^ of 0.984, and concordance rate of 0.988 in 4-digit allelic resolution. The differences in total accuracy were modest among the methods; however, DEEP*HLA was more advantageous for rare alleles (For alleles with a frequency < 1%, sensitivity = 0.690; PPV = 0.799; *r*^*2*^ = 0.911; and concordance rate = 0.691 in DEEP*HLA, compared to sensitivity = 0.628, 0.635; PPV = 0.624, 0.505; *r*^*2*^ = 0.862, 0.792; and concordance rate = 0.621, 0.675 in SNP2HLA and HIBAG, respectively; **Fig. 2a**). Further, we applied the model trained with our Japanese reference panel to a dataset of 908 Japanese individuals to investigate whether DEEP*HLA could impute well when applied to independent samples. The dataset comprised 4-digit alleles of 8 classical HLA genes based on the SSO method and SNP data genotyped using multiple genotyping arrays.^6^ DEEP*HLA achieved the highest total accuracy, with a sensitivity of 0.973, PPV of 0.972, *r*^*2*^ of 0.986, and concordance rate of 0.973. Again, DEEP*HLA was more advantageous for low-frequency and rare alleles (**Fig. 2a**). For alleles with a frequency < 1%, sensitivity = 0.690; PPV = 0.799; *r*^*2*^ = 0.911; and concordance rate = 0.691 in DEEP*HLA, compared to sensitivity 8 = 0.628, 0.635; PPV = 0.624, 0.505; *r*^*2*^ = 0.862, 0.792; and concordance rate = 0.621, 0.675 in SNP2HLA and HIBAG, respectively.

**Figure 2.**
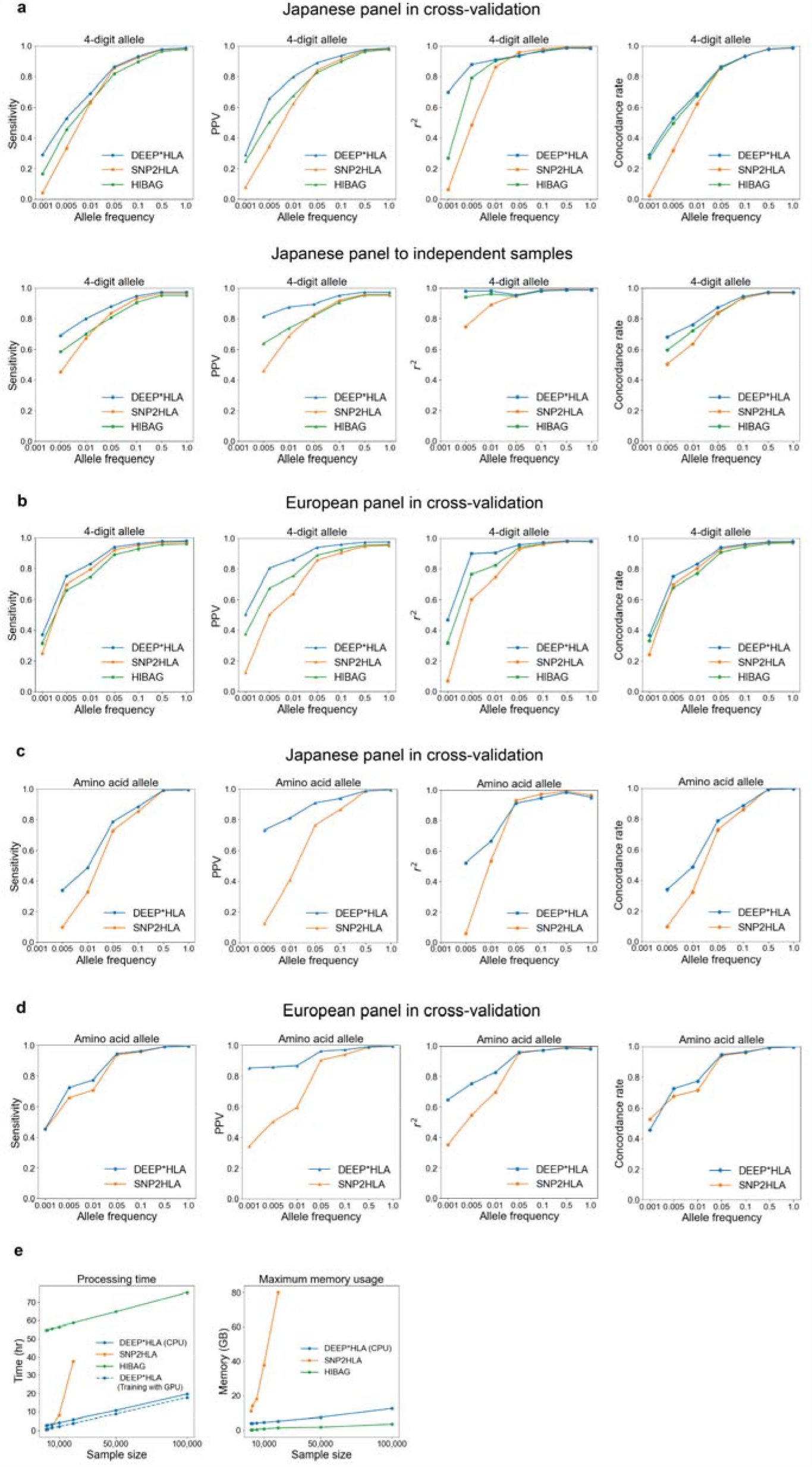
Performance evaluations of HLA imputation methods. (**a-d**) Sensitivity, PPV, and *r*^*2*^ of allelic dosage and concordance rate of best-guess genotypes for the 4-digit alleles (**a, b**) and amino acid polymorphisms (**c, d**) evaluated in our Japanese reference panel (**a, c**) and the T1DGC reference panel (**b, d**). For each metric, mean values of alleles with a frequency less than a value on the horizontal axis are shown on the vertical axis. DEEP*HLA was advantages especially for rare alleles. (**e**) Processing time (left) and maximum memory usage (right) evaluated on imputing BBJ samples using the Japanese panel. DEEP*HLA imputed by far the fastest in total processing time as the sample size increased. The dashed blue line in the processing time represents a case when DEEP*HLA used GPU only in training a model. All methods exhibited maximum memory usage scaling roughly linearly with sample size. SNP2HLA did not work within 100 GB in our machine for the sample sizes greater than 20,000.

We also applied DEEP*HLA to the Type 1 Diabetes Genetics Consortium (T1DGC) reference panel of 5,122 unrelated individuals of European ancestries.^33^ It comprises 2- and 4-digit alleles of the 8 classical HLA genes based on the SSO method, with SNP data of the MHC region genotyped with the Illumina Immunochip. DEEP*HLA achieved a sensitivity of 0.979, PPV of 0.976, *r*^*2*^ of 0.981, and concordance rate of 0.979 in 4-digit resolution, and these values were superior to those of SNP2HLA and HIBAG. DEEP*HLA was more advantageous especially in PPV and *r*^*2*^, for low-frequency and rare alleles (**Fig. 2b**). For alleles with a frequency < 1%, sensitivity = 0.830; PPV = 0.863; *r*^*2*^ = 0.908; and concordance rate = 0.832 in DEEP*HLA, compared to sensitivity = 0.793, 0.745; PPV = 0.640, 0.753; *r*^*2*^ = 0.745, 0.886; and concordance rate = 0.804, 0.769 in SNP2HLA and HIBAG, respectively.

We assessed the superiority of DEEP*HLA using a down-sampling approach (**Supplementary Note 1a**). DEEP*HLA trained with down-sampled data also outperformed other methods especially for rare allele, although there were differences between metrics (**Supplementary Fig. 1**). In the cross-validation of our Japanese reference panel, DEEP*HLA with sampling rates of 70%–80% and 60%–70% was almost equivalent to HIBAG and SNP2HLA, respectively. In the Japanese independent samples, DEEP*HLA with a sampling rate of even 70% and 60% outperformed HIBAG and SNP2HLA, respectively. In the cross-validation of the T1DGC panel, DEEP*HLA with a sampling rates of 70%–80% was almost equivalent to HIBAG and SNP2HLA, respectively. Notably, DEEP*HLA with a sampling rate of even 50% outperformed other methods in most cases in terms of PPV.

Finally, we investigated differences in accuracy among different HLA genes (**Fig. 3**). Whereas the accuracies for *HLA-B* and *HLA-DRB1* were lower than those for other loci especially in terms of total accuracy, those in DEEP*HLA were relatively high. As a result, DEEP*HLA had the highest means and lowest variances of accuracies among HLA genes in most cases. Only for rare alleles in the Japanese independent samples, the variances of sensitivity and concordance rate were higher than those for SNP2HLA, in which the accuracy metrics of SNP2HLA were lower than those of DEEP*HLA for almost all loci.

**Figure 3.**
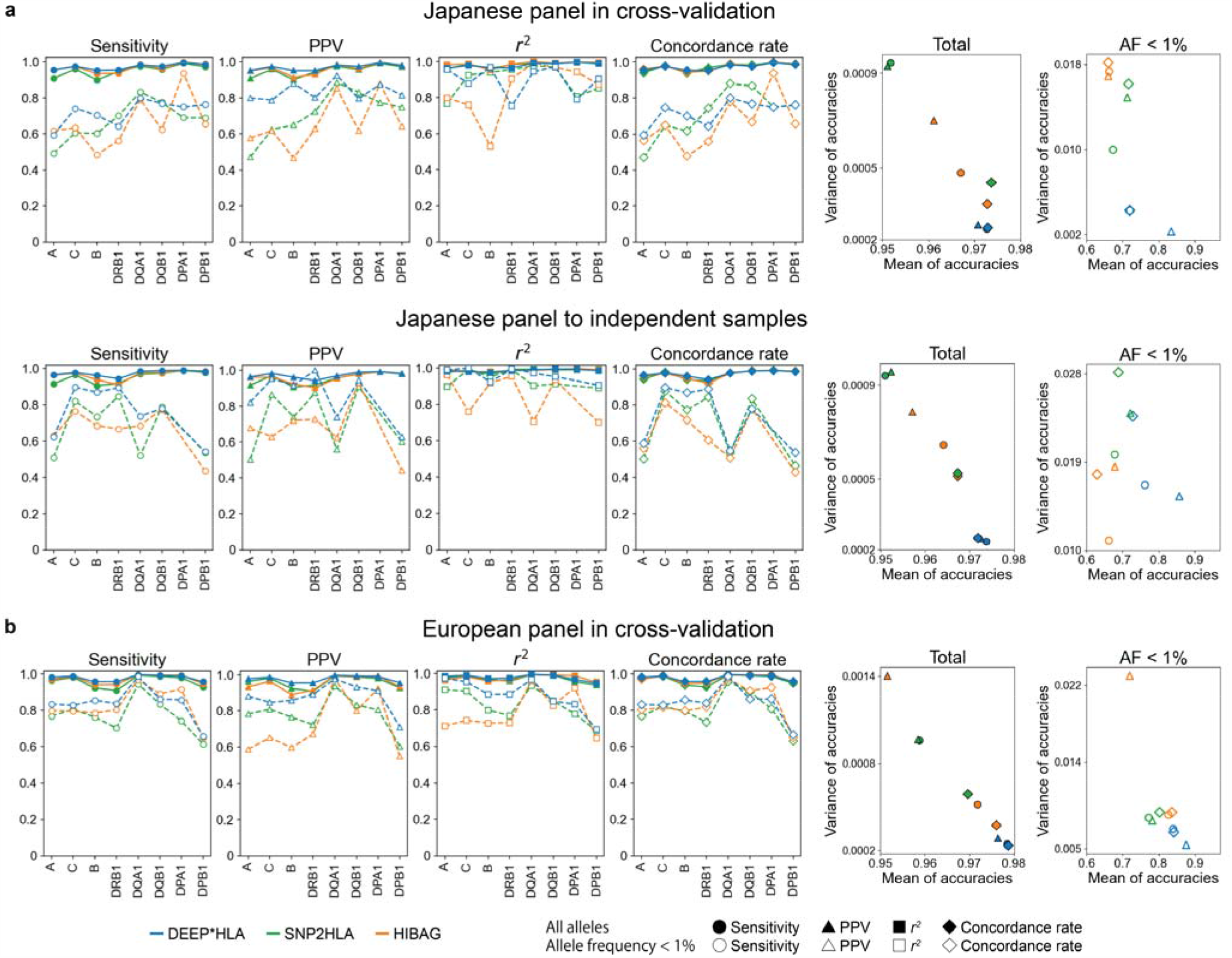
Comparison of imputation accuracy between different HLA genes. Each panel represents accuracy in 8 classical HLA genes evaluated in the Japanese panel in cross-validation (a, upper), the Japanese panel to the independent data (a, lower), and the European panel in cross-validation (b). Solid and dashed lines correspond to the accuracy of all allele and allele with frequency < 1%, respectively. The right two scatter plots represent the relation between the mean and variance of each metric among different HLA genes for individual methods. *R*^2^ metric is not shown because it is not an additive statistic.

In summary, although the improvement in total accuracy of DEEP*HLA might be modest, DEEP*HLA was advantageous in imputing infrequent alleles especially in terms of the dosage accuracy. PPV was significantly decreased in SNP2HLA, probably because the sum of the allele dosages of each HLA gene in an individual can exceed the expected value (i.e. = 2.0) since it imputes each allele separately as a binary allele. The improvement in dosage accuracy is meaningful considering that allelic dosages are typically used for association analysis.^3^ Furthermore, its small inter-locus variation in imputation accuracy should also be advantageous in MHC fine-mapping because the accuracy difference among HLA genes would result in imbalanced filtering, leading to a biased result.

### DEEP*HLA achieved higher accuracy when applied to 1000 Genomes Project data using a mixed reference panel

To conduct further validation in independent samples and evaluate the effect of ethnicity differences between a reference panel and target populations, we tested imputation accuracy in 1KGv3 cohort. First, we conducted HLA imputation using our Japanese panel and (*n* = 1,118) and a mixed panel which was experimentally conducted using the Japanese and the European panels (*n* = 6,240). When we used the Japanese panel, DEEP*HLA achieved the highest accuracies in all the metrics in the 1KGv3 JPT cohort (sensitivity = 0.974, PPV = 0.950, *r*^*2*^ = 0.995, and concordance rate = 0.975 in total alleles; **Supplementary Fig. 2a**). All the methods achieved high accuracies for rare alleles, in which DEEP*HLA was still superior (sensitivity = 0.862, PPV = 0.865, *r*^*2*^ = 0.999, and concordance rate = 0.862 for alleles with a frequency of < 1%). On the other hand, in other populations including EAS (excluding JPT), no methods were found to be accurate enough for practical use. This is probably attributed to the distinct haplotype structures and allele frequency spectra specific for Japanese ancestries even within East Asian populations.^6^ In addition, DEEP*HLA did not always perform better than other methods. Presumably, its high learning capacity of deep learning might backfire and caused overfitting to the population-specific reference panel. We thus recommend empirical validation of accuracy when applying DEEP*HLA to individuals mismatched with a reference panel population.

When we used a mixed panel, despite a slight decline in accuracy in JPT (sensitivity = 0.965, PPV = 0.940, *r*^*2*^ = 0.996, and concordance rate = 0.964 for total alleles), DEEP*HLA achieved high accuracies in EUR populations (sensitivity = 0.964, PPV = 0.918, *r*^*2*^ = 0.983, and concordance rate = 0.963 for total alleles). DEEP*HLA also achieved the highest accuracies in both JPT and EUR populations for total and rare alleles although the difference was relatively modest (**Supplementary Fig. 2b**). Thanks to a significant increase in the sample sizes of the reference panel, the accuracies in other populations were also improved. Notably, DEEP*HLA achieved the highest accuracies in the different populations, especially for rare alleles. Although the mixed panel used here is an experimental version that comprises genotypes from different typing procedures, the present results would suggest the applicability of our method to a multi-ethnic reference panel.

### DEEP*HLA can define HLA amino acid polymorphisms consistently with classical alleles

DEEP*HLA separately imputes classical alleles of each HLA gene, as a multi-class classification in the field of machine learning. Thus, it has an advantage that the sum of imputed allele dosages of each HLA gene is definitely set as an ideal value of 2.0. This enables us to define a dosage of amino acid polymorphisms from the imputed 4-digit allele dosages consistently with classical alleles. We compared this method of imputing amino acid polymorphisms with SNP2HLA, which imputes each allele as binary alleles. Although DEEP*HLA was equivalent to SNP2HLA in imputing amino acid polymorphisms in total alleles (sensitivity = 0.996, PPV = 0.996, *r*^*2*^ = 0.951, and concordance rate = 0.996 in the Japanese panel; sensitivity = 0.997, PPV = 0.995, *r*^*2*^ = 0.982, and concordance rate = 0.997 in T1DGC panel), it achieved more accurate imputation for rare alleles (sensitivity = 0.487, PPV = 0.811, *r*^*2*^ = 0.665, and concordance rate = 0.487 in the Japanese panel; sensitivity = 0.775, PPV = 0.864, *r*^*2*^ = 0.826, and concordance rate = 0.775 in T1DGC panel for alleles with a frequency of < 1%; **Fig. 2c, d**). The improvement in performance in terms of PPV was remarkable.

We admit that this method is only applicable to the reference panel where 4-digit alleles are accurately determined. Therefore, our method could not eliminate the ambiguity in the genotyping that derived from incompleteness of the original reference panel.

### High performance of DEEP*HLA in computational costs

We benchmarked the computational costs of DEEP*HLA against those of SNP2HLA and HIBAG using a subset of the GWAS dataset from BBJ containing *n* = 1,000, 2,000, 5,000, 12 10,000, 20,000, 50,000, and 100,000 samples (2,000 SNPs was consistent with the reference panel). A model-training process with reference data is required for DEEP*HLA and HIBAG but not for SNP2HLA. In addition, DEEP*HLA took an input of pre-phased GWAS data. Thus, we compared the total processing time including pre-phasing of GWAS data, model training, and imputation of DEEP*HLA, with the time of model training and imputation of HIBAG, and the running time of SNP2HLA. As shown in **Fig. 2e**, DEEP*HLA imputation had by far the fastest total processing time as the sample size increased. On comparing pure imputation times, it was faster than HIBAG (**Supplementary Table 1**). Furthermore, with a state-of-the-art GPU, the training time of DEEP*HLA was shortened from 153 min to 36 min. As for memory cost, all methods exhibited maximum memory usage scaling roughly linearly with sample size (**Fig. 2e** and **Supplementary Table 1**). HIBAG was the most memory-efficient across all sample sizes. Whereas SNP2HLA could not run within our machine’s 100 GB memory for sample sizes of >20,000, DEEP*HLA was able to perform imputation even for biobank-scale sample sizes of 100,000.

### Characteristics of the alleles for which DEEP*HLA was advantageous to impute

We focused on the characteristics of the HLA alleles of which accuracy was improved by DEEP*HLA compared with SNP2HLA, which is a gold-standard software. SNP2HLA runs Beagle intrinsically, which performs imputation based on a hidden Markov model of a localized haplotype-cluster. We hypothesized that this kind of methods shows better performance for imputing alleles for which LDs with the surrounding SNVs are stronger in close positions and get weaker as the distance from the target HLA allele increases (we termed this feature as distance-dependent LD decay). Conversely, it might show limited performance for imputing alleles with sparse LD structures throughout the MHC region. We defined the area under the curve (AUC) representing distance-dependent LD decay to verify this hypothesis. AUC values increase when LDs with the surrounding SNVs get stronger as they get closer to the target HLA allele (**Fig. 4b**). We evaluated the degree by which the accuracies of DEEP*HLA and SNP2HLA were affected by the AUCs and allele frequency using multivariate linear regression analysis. When calculating AUCs, we tested two different window sizes of AUCs: bilateral 1,000 SNPs from a target HLA allele and input size of DEEP*HLA. As expected, all accuracy metrics of SNP2HLA were positively correlated with the AUCs. Although the accuracy metrics of DEEP*HLA were also correlated with AUC, the correlations were weaker than those in SNP2HLA for all the metrics in both reference panels (**Fig. 4a** and **Supplementary Table 2**). In addition, we assessed the correlation between a simple metric of the maximum value of LD coefficients within 100 SNPs from a target allele and the accuracy of each method to examine our assumption more robustly with another index. Similarly, the correlations in DEEP*HLA were weaker than those in SNP2HLA (**Supplementary Table 2**).

**Figure 4.**
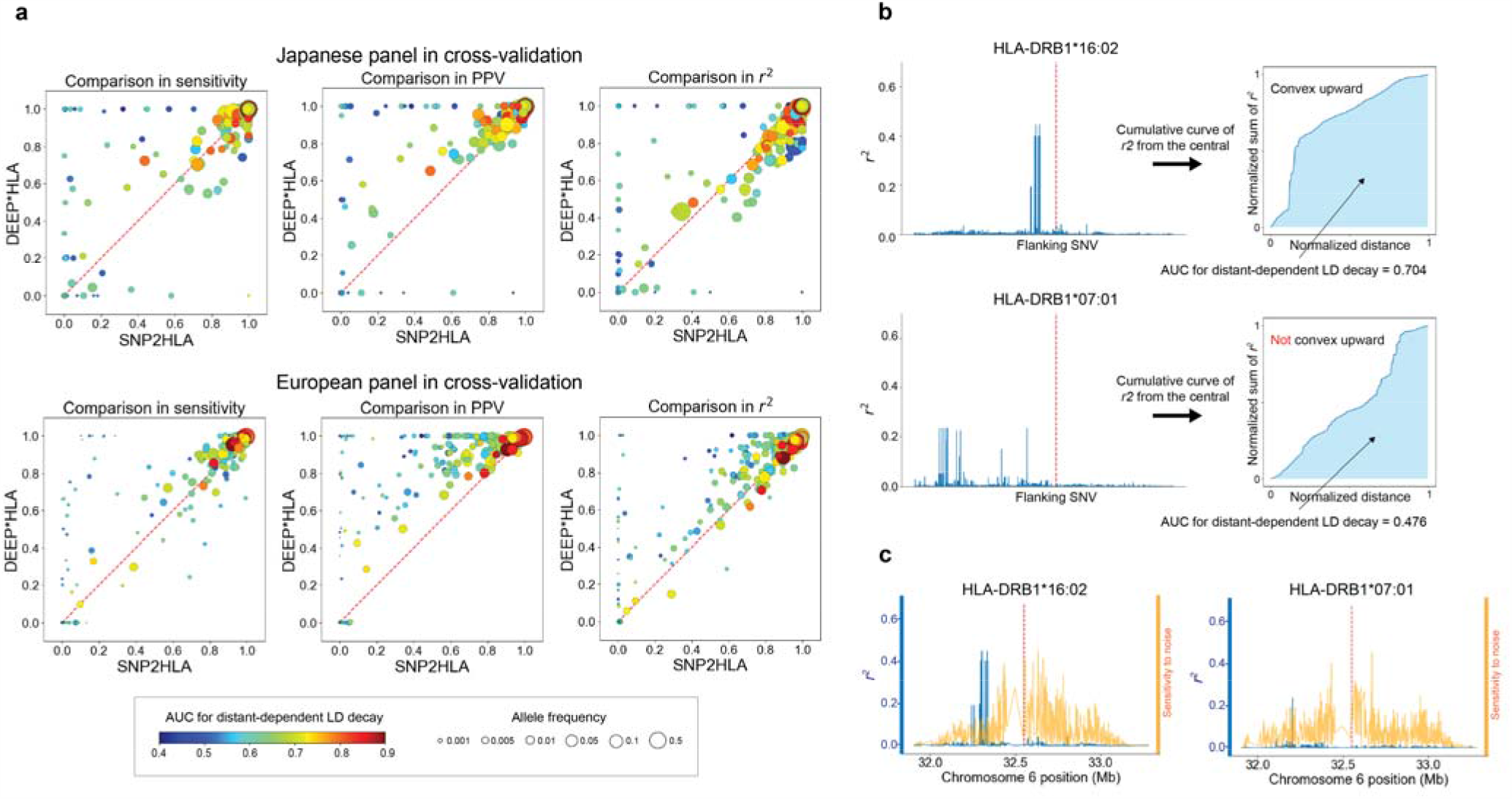
Comparison between DEEP*HLA and SNP2HLA displayed with allele frequencies and AUC for distance-dependent LD decay. (**a**) Comparisons of imputation accuracy between DEEP*HLA and SNP2HLA for 4-digit allele imputation for cross-validation with the Japanese panel (upper) and T1DGC panels (lower). Each dot corresponds to one allele, displayed with allele frequencies (size) and AUC for distance-dependent LD decay (color). The AUC was calculated based on bilateral 1,000 SNPs. Comparisons in concordance rate are not shown because they were almost the same as those in sensitivity. The performance of SNP2HLA was limited when imputing the alleles with low-frequency and low AUC; DEEP*HLA was relatively accurate even for the less frequent alleles regardless of AUC. (**b**) Example illustrations of AUC for distance-dependent LD decay. The left figures illustrate *r*^*2*^ of LD between an HLA allele (red dash line in the central) and flanking SNVs. HLA-DRB1*16:02 has strong LD in close positions and weaker LD in the distant positions. The cumulative curve of *r*^*2*^ of bilateral SNVs becomes convex upward; and the AUC increases. In contrast, HLA-DRB1*07:01 has moderate LD in distant or sparse positions, the curve does not become convex upward, and the AUC becomes smaller. (**c**) Comparison between *r*^*2*^ (blue line) and sensitivity maps of DEEP*HLA (orange line) for example alleles (red dash line in the center). The sensitivities are normalized for visibility. In both examples, DEEP*HLA reacted to noise across an extensive area regardless of LD.

Next, we used SmoothGrad to investigate our assumption that DEEP*HLA performs better imputation by recognizing distant SNVs as well as close SNVs of strong LD. SmoothGrad is a method for generating sensitivity maps of deep learning models.^34^ It is a simple approach based on the concept of adding noise to the input data and taking the mean of the resulting sensitivity maps for each sampled data. A trained DEEP*HLA model reacted to the noises of not only the surrounding SNVs with strong LD but also the distant SNVs as displayed in example HLA alleles (**Fig. 4c**). Interestingly, SNVs that reacted strongly were not always those of even moderate LD, but also spread across the entire the input region. While the validity of SmoothGrad for a deep learning model of genomic data is presently under investigation, one probable explanation is that predicting an allele using our method also means predicting the absence of other alleles of the target HLA gene. Thus, any SNV positions in LD with any of the other HLA alleles could be informative. Another explanation is that DEEP*HLA might recognize complex combinations of multiple distinct SNVs within the region rather than the simple LD correlations between HLA alleles and -SNVs.

### Empirical evaluation of imputation uncertainty

A common issue in deep learning models is quantification of the reliability of their predictions. One potential solution is uncertainty inferred from the concept of Bayesian deep learning.^35^ We experimentally evaluated imputation uncertainty by DEEP*HLA using Monte Carlo (MC) dropout, which could be applied following the general implementation of neural networks with dropout units.^36,37^ In MC dropout, uncertainty is presented as entropy of sampling variation with keeping dropout turned on. This uncertainty index corresponds not to each binary allele of a HLA gene, but to the prediction of genotype of each HLA gene of an individual. Thus, we evaluated whether the uncertainty could guess the correctness of best-guess genotypes of the target HLA genes. We compared this with a dosage-based discrimination, in which we assumed that a best-guess imputation of higher genotype dosage (probability) is more likely to be correct. The entropy-based uncertainty identified incorrectly-imputed genotypes with an areas under the curve of the receiver operating characteristic curve (ROC-AUC) of 0.851 in the Japanese panel and of 0.883 in the T1DGC reference panel in 4-digit alleles, which were superior to dosage-based discrimination (ROC-AUC = 0.722 and 0.754 in the Japanese T1DGC panels, respectively; **Supplementary Fig. 3**). Estimation of prediction uncertainty of a deep learning model is still under development;^37^ however, our results might illustrate its potential applicability to the establishment of a reliability score for genotype imputation by deep neural networks.

### Trans-ethnic MHC fine-mapping of T1D

We applied the DEEP*HLA models trained with our Japanese panel and the T1DGC panel to HLA imputation of T1D GWAS data from BBJ (831 cases and 61,556 controls) and UKB (732 cases and 353,727 controls), respectively. T1D is a highly heritable autoimmune disease that results from T cell–mediated destruction of insulin-producing pancreatic β cells.^38^ We performed imputation for GWAS data of the cohorts separately and then combined them to perform trans-ethnic MHC fine-mapping (1,563 cases and 415,283 controls). We filtered imputed alleles in which *r*^*2*^ accuracy in 10-fold cross-validation was lower than 0.7 in the current application.

Association analysis of the imputed HLA variants with T1D identified the most significant association at the HLA-DRβ1 amino acid position 71 (*P*_omnibus_ = *P* = 7.5 × 10^−120^; **Fig. 5a and Supplementary Table 3**), one of the T1D risk-associated amino acid polymorphisms in the European population.^12^ As for T1D, the largest HLA gene associations were reported for a combination of variants in the *HLA-DRB1, -DQA1*, and *-DQB1*;^12,39^ thus, we further investigated independently associated variants within these tightly linked HLA genes before searching for other risk-associated loci. When conditioning on HLA-DRβ1 amino acid position 71, we observed the most significant independent association in HLA-DQβ1 amino acid position 185 (*P*_omnibus_ = 3.1 × 10^−69^). Through stepwise forward conditional analysis in the class II HLA region, we found significant independent associations for Tyr30 in HLA-DQβ1 (*P*_binary_ = 6.7 × 10^−20^), HLA-DRβ1 amino acid position 74 (*P*_omnibus_ = 1.2 × 10^−11^), and Arg70 in HLA-DQβ1 (*P*_omnibus_ = 3.3 × 10^−9^; **Supplementary Fig. 4** and **Supplementary Table 4**).

**Figure 5.**
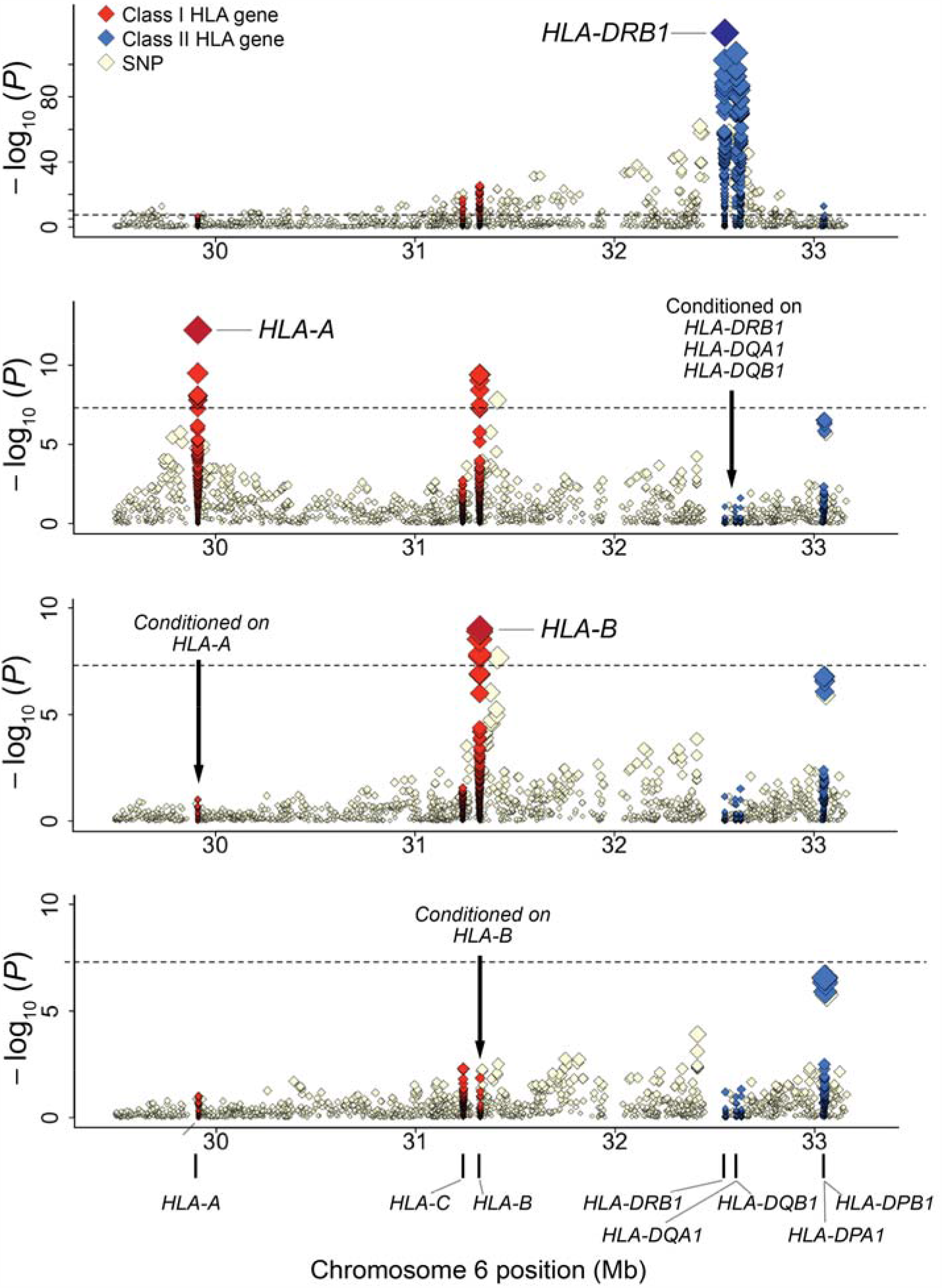
Trans-ethnic association plots of HLA variants with T1D in the MHC region. Diamonds represent‐log_10_ (*P* values) for the tested HLA variants, including SNPs, classical alleles, and amino acid polymorphisms of the HLA genes. Dashed black horizontal lines represent the genome-wide significance threshold of *P* = 5.0 × 10^−8^. The physical positions of the HLA genes on chromosome 6 are shown at the bottom. (**a–e**) Each panel shows the association plot in the process of stepwise conditional regression analysis: nominal results. (**a**) Results conditioned on *HLA-DRB1, HLA-DQA1*, and *HLA-DRB1*. (**b**) Results conditioned on *HLA-DRB1, HLA-DQA1, HLA-DRB1*, and *HLA-A*. (**c**) Results conditioned on *HLA-DRB1, HLA-DQA1, HLA-DRB1, HLA-A*, and *HLA-B*. (**d**) Our study identified the independent contribution of multiple HLA class I and class II genes to the T1D risk in a trans-ethnic cohort, in which the impacts of class II HLA genes were more evident. Detailed association results are shown in **Supplementary Table 3**.

These results were different from those of a previous study of a large T1D cohort of European ancestries, which reported three amino acid polymorphisms, i.e., HLA-DQβ1 position 57, HLA-DRβ1 position 13, and HLA-DRβ1 position 71, as the top-associated amino acid polymorphisms in the *HLA-DRB1, -DQA1*, and *-DQB1* region. We then constructed multivariate regression models for individual populations that incorporated our T1D risk-associated HLA amino acid polymorphisms and classical alleles of *HLA-DRB1* and *HLA-DQB1*, and compared the effects of these variants. The odds ratios of the risk-associated variants reported previously did not show any positive correlation between different populations (Pearson’s *r* =‐0.59, *P* = 0.058; **Supplementary Fig. 5** and **Supplementary Table 5**). On the other hand, we identified a set of variants with significant positive correlation by trans-ethnic fine-mapping of the integrated cohort data (Pearson’s *r* = 0.76, *P* = 6.8 × 10^−3^; **Supplementary Fig. 5**).

We further investigated whether the T1D risk was associated with other HLA genes independently of *HLA-DRB1, -DQA1*, and *-DQB1*. When conditioning on *HLA-DRB1, -DQA1*, and *-DQB1*, we identified a significant independent association at HLA-A amino acid position 62 (*P*_omnibus_ = 5.9 × 10^−13^; **Fig. 5b** and **Supplementary Table 3**). After conditioning on HLA-A amino acid position 62, we did not observe any additional independent association in HLA-A alleles. When we conditioned on *HLA-DRB1, -DQA1, -DQB1*, and *-A*, we identified a significant independent association at HLA-B*54:01 (*P*_binary_ = 1.3 × 10^−9^; **Fig. 5c** and **Supplementary Table 8**), and its unique amino acid polymorphisms (Gly45 and Val52 at HLA-B). When conditioned on *HLA-DRB1, -DQA1, -DQB1, -A*, and *-B*, no variants in the MHC region satisfied the genome-wide significance threshold (*P* > 5.0 × 10^−8^; **Fig. 5d** and **Supplementary Table 3**). Multivariate regression analysis of the identified risk variants explained 10.3% and 27.6% of the phenotypic variance in T1D under assumption of disease prevalence of 0.014%^40^ and 0.4%.^41^ for the Japanese and British cohorts, respectively. Their odds ratios on T1D risk were also correlated between different populations (Pearson’s *r* = 0.71, *P* = 4.4 × 10^−3^; **Fig. 6** and **Table 1**).

**Figure 6.**
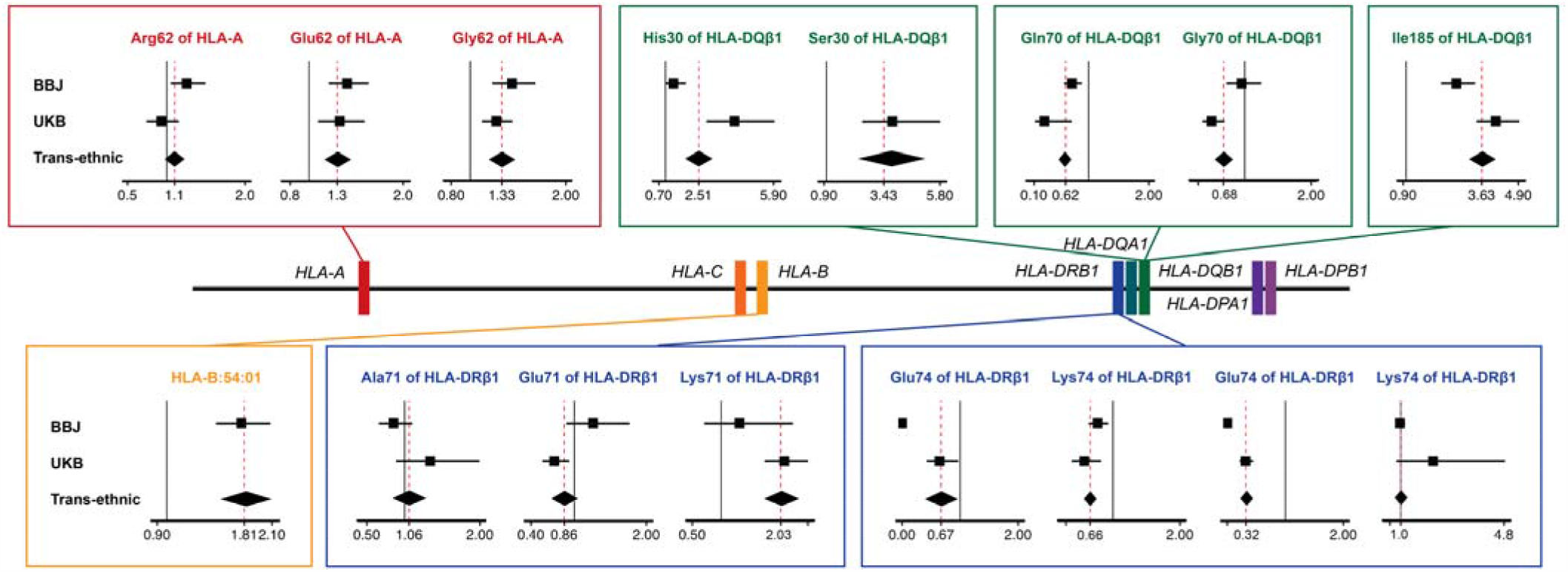
HLA variants associated with the T1D risk identified through trans-ethnic fine-mapping. Forest plots for individual risk-associated alleles are displayed along with a location map of classical HLA genes. Each forest plot shows the estimated odds ratio (OR) and 95% confidence interval from cohort-specific logistic model for BBJ and UKB, and the trans-ethnic logistic model. Red dashed lines indicate OR in trans-ethnic cohorts. Black solid lines represent OR = 1. Colored square boxes represent amino acid polymorphisms of the same position or a classical allele.

**Tables 1.** Associations of the HLA variants with the T1D risk identified through trans-ethnic fine-mapping study.

| HLA variant | Frequency (BBJ) |  | Frequency (UKB) |  | OR (95% CI) |  | P† |  |
| --- | --- | --- | --- | --- | --- | --- | --- | --- |
|  | Case<br>n = 831 | Control<br>n = 61,556 | Case<br>n = 732 | Control<br>n = 353,727 | BBJ | UKB | BBJ | UKB |
| HLA-DRβ1 amino acid position 71 |  |  |  |  |  |  |  |  |
| Alanine | 0.10 | 0.18 | 0.04 | 0.15 | 0.85 (0.66-1.10) | 1.34 (0.89-1.99) | 0.23 | 0.16 |
| Arginine | 0.82 | 0.73 | 0.33 | 0.45 | (reference) |  |  |  |
| Glutamic acid | 0.073 | 0.074 | 0.083 | 0.12 | 1.26 (0.89-1.77) | 0.72 (0.56-0.93) | 0.019 | 0.0013 |
| Lysine | 0.0096 | 0.011 | 0.54 | 0.28 | 1.31 (0.71-2.24) | 2.11 (1.77-2.53) | 0.035 | 1.9 × 10<sup>-16</sup> |
| HLA-DQβ1 amino acid position 185 |  |  |  |  |  |  |  |  |
| Isoleucine | 0.39 | 0.57 | 0.68 | 0.83 | 2.74 (2.21-3.40) | 4.12 (3.49-4.99) | 3.5 × 10<sup>-20</sup> | 7.0 × 10<sup>-55</sup> |
| Threonine | 0.61 | 0.43 | 0.32 | 0.17 | (reference) |  |  |  |
| HLA-DQβ1 amino acid position 30 |  |  |  |  |  |  |  |  |
| Histidine | 0.16 | 0.19 | 0.18 | 0.23 | 1.36 (0.97-1.93) | 4.16 (2.86-5.96) | 0.0078 | 3.0 × 10<sup>-14</sup> |
| Serine | 0.0042 | 0.0038 | 0.34 | 0.25 | inf | 3.82 (2.53-5.87) | 0.079 | 3.8 × 10<sup>-10</sup> |
| Tyrosine | 0.83 | 0.80 | 0.48 | 0.52 | (reference) |  |  |  |
| HLA-DRβ1 amino acid position 74 |  |  |  |  |  |  |  |  |
| Alanine | 0.56 | 0.59 | 0.59 | 0.65 | (reference) |  |  |  |
| Arginine | 0.0018 | 0.00088 | 0.28 | 0.15 | 0 (0-0.045) | 0.64 (0.42-0.96) | 0.08 | 0.0036 |
| Glutamic acid | 0.32 | 0.27 | 0.021 | 0.036 | 0.77 (0.64-0.93) | 0.57 (0.38-0.82) | 0.00065 | 0.0004 |
| Glutamine | 0.0024 | 0.0030 | 0.0795 | 0.15 | 0 (0-0.0029) | 0.31 (0.21-0.44) | 0.079 | 4.5 × 10<sup>-10</sup> |
| Leucine | 0.12 | 0.14 | 0.023 | 0.023 | 0.97 (0.81-1.16) | 2.20 (0.85-4.84) | 0.074 | 0.0077 |
| HLA-DQβ1 amino acid position 70 |  |  |  |  |  |  |  |  |
| Arginine | 0.60 | 0.62 | 0.79 | 0.63 | (reference) |  |  |  |
| Glutamic acid | 0.26 | 0.17 | 0.020 | 0.020 | 0.73 (0.59-0.9) | 0.27 (0.11-0.71) | 0.00020 | 0.0052 |
| Glycine | 0.14 | 0.20 | 0.19 | 0.35 | 0.95 (0.72-1.25) | 0.50 (0.36-0.70) | 0.073 | 3.1 × 10<sup>-5</sup> |
| HLA-A amino acid position 62 |  |  |  |  |  |  |  |  |
| Arginine | 0.19 | 0.20 | 0.06 | 0.09 | 1.25 (1.05-1.49) | 0.93 (0.74-1.16) | 0.0012 | 0.53 |
| Glutamic acid | 0.39 | 0.37 | 0.09 | 0.09 | 1.40 (1.21-1.63) | 1.33 (1.10-1.60) | 9.2 × 10<sup>-6</sup> | 0.0025 |
| Glutamine | 0.15 | 0.19 | 0.46 | 0.49 | (reference) |  |  |  |
| Glycine | 0.26 | 0.24 | 0.33 | 0.29 | 1.44 (1.23-1.68) | 1.27 (1.12-1.44) | 6.6 × 10<sup>-6</sup> | 1.6 × 10<sup>-4</sup> |
| Leucine | 0 | 0 | 0.055 | 0.044 | - | 2.01 (1.57-2.55) | 1.5 × 10<sup>-12</sup> | 1.9 × 10<sup>-8</sup> |
| HLA-B*54:01 | 0.14 | 0.073 | 0 | 0 | 1.78 (1.51-2.08) | - | - | - |

HLA, human leucocyte antigen; OR, odds ratio; 95% CI, 95% confidence interval.
†Obtained from the multivariate regression model that included all the variants listed here.

To evaluate the advantage of the trans-ethnic fine-mapping, we performed fine-mapping for each cohort separately and compared the results with those of the trans-ethnic analysis. The most significant associations were observed in the *HLA-DRB1* and *-DQB1* in bot h cohorts (**Supplementary Fig. 6** and **Supplementary Fig. 7**). The top signals were at the HLA-DQβ1 amino acid position 185 (*P* = 8.3 × 10^−47^) for the BBJ cohort and HLA-DRβ1 amino acid position 71 (*P* = 4.1 × 10^−107^) for the UKB cohort, both of which were consistent with the risk-associated variants identified through the trans-ethnic fine-mapping. On the other hand, the risk-associated variants pointed in subsequent conditional analyses within this region were not identical. Generally, parsimonious fine-mapping using a single population was challenging due to multiple candidate variants with similar degrees of LD (and thus associations) to the top signal in each iteration of the stepwise conditional analysis (**Supplementary Fig. 8** and **Supplementary Fig. 9**). As a result of the trans-ethnic analysis, we successfully identified finer sets of the more variants, which exhibited clearer significance by interrogating the different LD patterns between the populations. When conditioning on *HLA-DRB1, -DQA1*, and *-DQB1*, we identified significant independent associations in *HLA-B* for the BBJ cohort with the top at HLA-B*54:01 (*P* = 4.1 × 10^−10^), and *HLA-A* for the UKB cohort with the top at HLA-A amino acid position 62 (*P* = 1.4 × 10^−8^), respectively (**Supplementary Fig. 6** and **Supplementary Fig. 7**). Both variants were identical to those originally identified in the trans-ethnic analysis. This observation indicates that the trans-ethnic analysis could discover more associated loci than single population-based analyses. Whereas HLA-B*54:01 was too rare and not assessed in Europeans, it is notable that the T1D risk of HLA-A amino acid position 62 was shared with East Asians. These observations should illustrate the value of the trans-ethnic MHC fine-mapping.

## Discussion

In this study, we demonstrated that DEEP*HLA, a multi-task convolutional deep learning method for HLA imputation, outperformed conventional HLA imputation methods in various aspects. DEEP*HLA was more advantageous when the target HLA variants, including classical alleles and amino acid polymorphisms, were low-frequent or rare. Our study demonstrated that the performance of a conventional method was reduced for alleles that did not exhibit distance-dependent LD decay with the target HLA allele. DEEP*HLA was less dependent on this point, and might comprehensively capture the relationships among multiple distinct variants regardless of LD. Taking advantage of the significant improvement of imputation accuracy in rare alleles, we conducted trans-ethnic MHC fine-mapping of T1D. This approach could be performed as well using the conventional HLA imputation methods. However, the results obtained using DEEP*HLA should be more reliable because there were several risk-associated alleles which were rare only in one population.

To date, technical application of deep neural networks to population genetics data has been limited. In a previous attempt for genotype imputation, a sparse convolutional denoising autoencoder was only compared with reference-free methods.^32^ There might be two possible explanations for the success of our DEEP*HLA. First unlike genotype imputation by denoising autoencoders, which assumes various positions of missing genotypes in a reference panel to impute, the prediction targets were fixed to the HLA allele genotypes as a classification problem. Second, convolutional neural networks, which leverage a convolutional kernel that is capable of learning various local patterns, might be better suited for learning the complex LD structures in the MHC region.

We filtered alleles with poor imputation quality based on the results of cross-validation in the current application; however, an indicator of reliability could be further utilized. We demonstrated that the prediction uncertainty inferred from a Bayesian deep learning method had potential capability of identifying incorrectly-imputed alleles in a per-gene level. Our future work should establish a method to quantify per-allele imputation uncertainty that can be practically used as a filtering threshold for subsequent analyses.

As for the genetic features of the MHC region associated with T1D, the highest risk is conferred by DR3-DQA1*05-DQB1*02 and DR4-DQA1*03-DQB1*03:02 haplotypes in Europeans,^39,42^ and by DR9-DQA1*03-DQB1*03:03 and DR4-DQA1*03-DQB1*04:01 haplotypes in Japanese.^43^ In a previous study for a large European cohort, Hu et al. demonstrated that the three amino acid polymorphisms of DRβ1 and HLA-DQβ1 explained the majority of the risk in the *HLA-DRB1, -DQA1*, and *-DQB1* region with the top signal at non-Asp57 in HLA-DQβ1.^12^ Conversely, the risk haplotypes in Japanese population carry Asp57 of HLA-DQβ1.^43^ We obtained several additional insights in the present study. We initially conducted a trans-ethnic MHC fine-mapping of T1D, and successfully disentangled a set of 5 risk-associated amino acid polymorphisms of position 71 and 74 in HLA-DRβ1, and 30, 70, and 185 in HLA-DQβ1. Four of these positions compose the peptide-binding grooves, suggesting their functional contributions to antigen-presentation ability (**Supplementary Fig. 10**). While the association of HLA-DRβ1 amino acid position 71 was replicated in concordance direction with Europeans, the effects in the Japanese population were not preserved in the final model. Whereas the association of amino acid position 74 in HLA-DRβ1 has been reported in Han-Chinese and certain European populations,^44,45^ the European study did not report its independent association due to the rareness of its characterized classical allele, HLA-DRB1*04:03. We successfully identified its independent association in trans-ethnic cohorts with a similar effect size between the diverse populations. Although amino acid position 185 in HLA-DQβ1 does not compose the peptide-binding groove, the variation of Ile/Thr is suggested to alter DQ-DM anchoring by interacting with its neighboring residues, leading to the susceptibility to other autoimmune diseases.^46,47^ Variant Ile185 is tagged with HLA-DQA1*03, which composes the risk haplotypes in Japanese and European population respectively. A correspondence table of the amino acid polymorphisms and 4-digit classical HLA alleles is shown in **Supplementary Table 6**. As a result, the catalogue of the T1D risk-associated variants in this region identified by our trans-ethnic approach was different from that in the European study.^12^ We admit the possibility that the smaller sample size in our study and different definitions of the phenotypes (between studies, and between cohorts in our study) might contribute to this disparity. Particularly, we note the potential distinctiveness of Japanese T1D phenotypes.^48^ However, considering that our observed variants shared the effects on the T1D risk between different populations, we might gain a novel insight into the issue of inter-ethnic heterogeneity of T1D risk alleles in the MHC region. As for class I HLA genes, the independent association of amino acid position 62 in HLA-A was consistent with the previous European study.^12^ We found that it had similar effects on the T1D risk also in the Japanese population. HLA-B*54:01 has traditionally been suggested as a potential risk allele in Japanese by a candidate HLA gene approach,^13^ of which an independent association via the MHC region-wide fine-mapping was first proven here.

While an advantage of trans-ethnic fine-mapping is the elucidation of truly risk-associated signals by adjusting confound by LD of each population,^49^ there are several potential limitations to note. First, we need to consider population-specific LD structures and allele frequency spectra, which are important especially in the MHC region. Strong population-specificity may preclude removal of the effects of LD for the current purpose of trans-ethnic fine-mapping when few populations are available. Conversely, some HLA alleles exist only in a certain population, and fine-mapping in a single population could also be of importance. Second, modeling heterogeneity in effects among diverse populations could enhance the power of discovery of causal variants in trans-ethnic analysis.^50^ Since the purpose of the current trans-ethnic fine-mapping is to identify trans-ethnically risk-associated variants rather than to discover variants with a strong effect only in one population, we did not explicitly model heterogeneity. However, in an analysis using more cohorts from different populations, modeling heterogeneity might be more suitable because a bias by single population would be reduced.

Therefore, multi-ethnic MHC fine-mapping that integrates further diverse ancestry should be warranted for robust prioritization of risk-associated HLA variants as a next step.^15^ Given their high learning capacity of deep neural networks, our method will be helpful not only when integrating the imputation results from multiple references, but also when using a more comprehensive multi-ethnic reference. We expect that highly accurate imputation realized by learning of complex LDs in the MHC region using neural networks will enable us to further elucidate the involvement of common genetic features in the MHC region that affect human complex traits across ethnicities.

## Data Availability

The Japanese HLA data have been deposited at the National Bioscience Database Center (NBDC) Human Database (research ID: hum0114). Independent HLA genotype data of Japanese population is available in the Japanese Genotype-phenotype archive (JGA; accession ID: JGAS00000000018). T1DGC HLA reference panel can be download at a NIDDK central repository with a request (https://repository.niddk.nih.gov/studies/t1dgc-special/). GWAS data of the BBJ are available at the NBDC Human Database (research ID: hum0014). UKBB GWAS data is available upon request (https://www.ukbiobank.ac.uk/).

## Acknowledgements

We would like to thank all the participants involvement in this study. We thank the members of Biobank Japan and RIKEN Center for Integrative Medical Sciences for their supports on this study.

## Data availability

The Japanese HLA data have been deposited at the National Bioscience Database Center (NBDC) Human Database (research ID: hum0114). Independent HLA genotype data of Japanese population is available in the Japanese Genotype-phenotype archive (JGA; accession ID: JGAS00000000018). T1DGC HLA reference panel can be download at a NIDDK central repository with a request (https://repository.niddk.nih.gov/studies/t1dgc-special/). GWAS data of the BBJ are available at the NBDC Human Database (research ID: hum0014). The analysis of UKB GWAS data was conducted via the application number 47821 (https:www.ukbiobank.ac.uk/).

## Code availability

Python scripts for training a model and performing imputation with our method are in DEEP*HLA GitHub repository (https://github.com/tatsuhikonaito/DEEP-HLA).

## Methods

### The architecture of DEEP*HLA

DEEP*HLA is a multi-task convolutional neural network comprising a shared part of two convolutional layers and a fully-connected layer, and individual fully-connected layers that output allelic dosages of individual HLA genes to simultaneously impute HLA genes of the same group (**Fig. 1a**). The grouping was based on the LD structure^3^ and physical distance in the current application: (1) {*HLA-F, HLA-V, HLA-G, HLA-H, HLA-K, HLA-A, HLA-J, HLA-L*, and *HLA-E*}, (2) {*HLA-C, HLA-B, MICA*, and *MICB*}, (3) {*HLA-DRA, HLA-DRB9, HLA-DRB5, HLA-DRB4, HLA-DRB3, HLA-DRB8, HLA-DRB7, HLA-DRB6, HLA-DRB2, HLA-DRB1, HLA-DQA1, HLA-DOB*, and *HLA-DQB1*}, and (4) {*TAP2, TAP1, HLA-DMB, HLA-DMA, HLA-DOA, HLA-DPA1*, and *HLA-DPB1*}. Genes not typed or with only single alleles in individual reference panels were excluded from the group. Comparisons with single-task neural networks or multi-task neural networks with random groupings are shown in **Supplementary Note 1b** and **Supplementary Fig. 11**.

DEEP*HLA takes the input of each haplotype SNV genotypes from pre-phased data, and outputs the genotype dosages of individual alleles for each HLA gene. For each group, SNVs within its window are encoded to one-hot vectors based on whether each genotype is consistent with a reference or alternative allele. The window sizes on each side were fixed to 500 kb for fair comparisons in the current investigation; using different window sizes might slightly change the accuracy for some loci (**Supplementary Note 1c** and **Supplementary Fig. 12**). Two convolutional layers with max-pooling layers and a fully-connected layer follow the input layer as a shared part. The fully-connected layer at the end of the shared part is followed by individual fully-connected layers which have nodes consistent with the number of alleles of each HLA gene. Softmax activation was added before the last output to return an imputation dosage that ranges from 0.0 to 1.0 for each allele of one haplotype. Thus, an individual layer outputs the individual allelic dosages of the HLA gene of which the sum equals 1 for one haplotype. Dropout was used on the convolutional and fully-connected layers,^51^ and batch normalization was added to the convolutional layers.^52^

During training, 5% of the data set were used for sub-validation to determine the point for early-stopping training. In 10-fold cross-validation, we separated sub-validation for early-stopping from a training fold to conduct valid benchmarking (**Supplementary Fig. 13**). A categorical cross entropy loss function for each HLA gene was minimized using the Adam optimizing algorithm.^53^ For a multi-task learning to find a Pareto optimal solution of all tasks, we used the multiple-gradient descent algorithm – upper bound (MGDA-UB), where the loss function of each task was scaled based on its optimization algorithms.^54^ To taking advantage of the hierarchical nature of HLA alleles (i.e. 2-digit, 4-digit, and 6-digit), we implemented hierarchical fine-tuning, in which parameters of the model of upper hierarchical structures were transferred to those of the lower one.^55^ We transferred the parameters of shared networks of 2-digit alleles to 4-digit alleles, and of 4-digit alleles to 6-digit alleles successively during training. Although some HLA alleles in our reference panel were not determined in 4-digit or 6-digit resolution, we set their upper resolution instead to maintain equivalent hierarchical levels with other HLA genes. Hyperparameters, including the number of filters and kernel sizes of convolutional layers, and fully-connected layer size, were tuned using Optuna.^56^ The hyperparameters for each reference panel were determined using a randomly sampled dataset before cross-validation. Our deep learning architectures were implemented using Pytorch 1.4.1 (see URLs), a Python neural network library.

### Empirical evaluation of HLA imputation accuracy

We used the accuracy metrics of sensitivity, PPV, and *r*^*2*^ for imputed allelic dosage, and concordance rate for best-guess genotypes to evaluate the imputation accuracy in various aspects.

In the paper of SNP2HLA, per-locus accuracy was defined as a sum of the dosage of each true allele across all individuals divided by the total number of observations.^33^ This definition of accuracy counts positives that are correctly identified as such and it corresponds to sensitivity in a cross-tabulation table when decomposed to individual alleles (**Supplementary Note 2** and **Supplementary Fig. 14**). Thus, we termed this as sensitivity (*Se*) to contrast with the PPV defined later

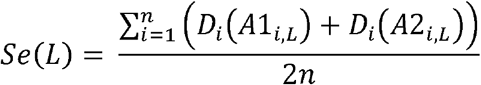

where *n* denotes the number of individuals, *D*_*i*_ represents the imputed dosage of an allele in individual *i*, and alleles *A1*_*i, L*_ and *A2*_*i, L*_ represent the true HLA alleles for individual *i* at locus *L*. The calculations were based on the condition that the imputed alleles are arranged to optimize for consistency with the truth alleles *A1*_*i, L*_ and *A2*_*i, L*_.

To evaluate the imputation performance in individual HLA alleles, we decomposed the *Se* (*L*) to evaluate the imputation performance of each allele as.

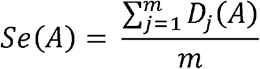

This metric cannot evaluate the effect of false positives; thus, we defined PPV in the same manner as

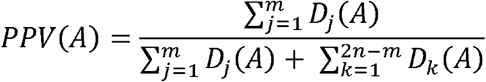

where *m* denotes the number of true observations of allele *A* in the total sample, and *D*_*i*_ represents imputed dosage of allele *A* in individual haplotype *j* that has allele *A. D*_*k*_ represents imputed dosage of allele *A* in individual haplotype *k* that has an allele other than allele *A*. This definition is also based on a cross-tabulation table (**Supplementary Fig. 14a**).

In addition, we calculated *r*^*2*^ based on Pearson’s product moment correlation coefficient between imputed and typed dosages for each allele.^22^

Further, to evaluate the accuracy of best-guess genotypes, we calculated the concordance rate (*CR*) of best-guess genotypes and true genotypes for each allele as

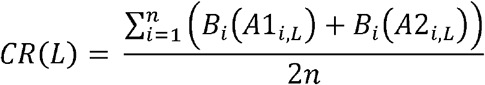

where *B*_*i*_ represents the best-guess genotype of an allele in individual *i*. By definition, it was the same as the sensitivity, in which dosages were changed to best-guess genotypes. Thus, we decomposed it to *CR*(*A*) for accuracy for each allele in the same way. We did not evaluate PPV for best-guess genotype due to redundancy.

When determining accuracy metrics for each locus or a certain range of allele frequencies, we calculated the weighted-mean of individual allele-level accuracies based on individual allele frequencies. For *r*^*2*^, we applied Fisher’s Z transformation to individual values, and back-transformed them after averaging to reduce bias.^57^

### Estimation of HLA imputation uncertainty of DEEP*HLA using MC dropout method

In order to estimate prediction uncertainty, we adopted the entropy of sampling variation of MC dropout method.^36^ In MC dropout, dropouts are kept during prediction to perform multiple model calls. Different units are dropped across different model calls; thus, it can be considered as Bayesian sampling with treating the parameters of a CNN model as random variables of Bernoulli distribution. The uncertainty of a best-guess genotype inferred from the entropy of sampling variation is determined as

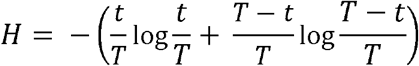

where *T* is the number of variational samplings and *t* is the number of times in which obtained genotype was identical to the best-guess genotype. We set *T* = 200 in the current investigation.

### AUC metric representing distance-dependent LD decay

To evaluate whether the the strength of LD between an HLA allele and its surrounding SNVs weakens as the the distance between them increases, we calculated the AUC of the cumulative curve of *r*^*2*^ from the HLA allele (AUC for distance-dependent LD decay). When the LD of flanking SNVs of an HLA allele has such a characteristic, *r*^*2*^ of LD from the HLA allele tends to decrease. In other words, the bilateral cumulative curve of *r*^*2*^ from the HLA allele is more likely to be convex upward; then, the AUC tends to be higher. We determined the AUC by normalizing the maximum values of *r*^*2*^ sum and window sizes to 1. We evaluated the association of the AUC with allele-level accuracy metrics of each imputation method by linear regression models adjusted for an allele frequency. The window size of the AUC should be set to an input range for each imputation method. However, SNP2HLA does not have a clear input range. Thus, we tested two different window sizes as bilateral 1,000 SNPs from a target HLA allele and the input range of DEEP*HLA. We investigated the correlation between the imputation accuracy and the AUC of two different window sizes, respectively.

### Regional sensitivity maps of DEEP*HLA

We applied SmoothGrad to estimate which SNVs were important for DEEP*HLA imputation of each HLA allele.^34^ For each haplotype, we generated 200 samples which were added Gaussian noise to encoded SNV data and input them into a trained model. Sensitivity values for individual SNV positions were obtained by averaging the absolute values of gradients caused by the difference from the true label. When we obtained the sensitivity of an HLA allele, we averaged the maps of all haplotypes that have the target HLA allele.

### HLA imputation software and parameter settings

We tested the latest version of the software available in Jun 2020 for comparison with our method. SNP2HLA (v1.0.3) first arranges the strand in its own algorithm; however, we removed this step data during cross-validation because the strands must be the same between training and test data. The other settings of SNP2HLA were set to the default values. For HIBAG (1.22.0.) the number of classifiers was set to 25, which is sufficient to achieve good performance^58^ for testing the Japanese data. For the T1DGC panel, the training time was extremely long with 25 classifiers; thus, we set 2 classifiers after we confirmed that the imputation accuracy was almost unchanged in the first set of cross-validation. The flanking regions on each side were set to 500 kb. The current version of HLA*IMP:02 did not support a function to generate an imputation model using own reference data in a publicly available form; thus, we did not evaluate its performance in this study for fair comparison.

### Measurement of computational costs

We measured the computational costs of imputation of a subset of BioBank Japan (BBJ) Project data set (*n* = 1,000, 2,000, 5,000, 10,000, 20,000, 50,000, and 100,000 samples) using our Japanese reference panel (2,000 SNVs were consistent). All our runtime analyses were performed on a dedicated server running CentOS 7.2.1511, with 48 CPU cores (Intel ® Xeon ® E5-2687W v4 @ 3.00 GHz) and 256 GB of RAM without GPU. Additionally, we measured the training time of DEEP*HLA with GPU using a machine with Ubuntu 16.04.6 LTS with 20 CPU cores (Intel ® Core (tm) i9-9900X @ 3.50 GHz), 2 GPUs (NVIDIA ® GeForce ® RTX 2080 Ti), and 128 GB of RAM. DEEP*HLA requires pre-phased GWAS data and the models trained with reference data; thus, we measured the process of not only imputation, but also pre-phasing of GWAS data (conducted by Eagle) and training the models with a reference panel. Similarly, HIBAG requires the time for training a model, which was also measured. In SNP2HLA, the maximum of available memory was set to 100 GB. The processing time and maximum memory usage were measured using GNU Time software when running from a command line interface.

### HLA imputation reference data

We used two HLA reference panels in cross-validation and HLA imputation for biobank GWAS data. The panels were distributed as a phased condition; thus, they were used as input for training a DEEP*HLA model as they were. When they were used as a validation set, we removed the target alleles (i.e. HLA alleles and amino acid alleles) to leave only phased SNP data. We discussed stricter cross-validation including the process of haplotype pre-phasing in **Supplementary Note 1d**.

#### (i) Our Japanese reference panel and a validation dataset

Our Japanese reference panel contained NGS-based 6-digit resolution HLA typing data of 33 classical and non-classical HLA genes, of which 9 were classical HLA genes (*HLA-A, HLA-B*, and *HLA-C* for class I; *HLA-DRA, HLA-DRB1, HLA-DQA1, HLA-DQB1, HLA-DPA1*, and *HLA-DPB1* for class II) and 24 were non-classical HLA genes (*HLA-E, HLA-F, HLA-G, HLA-H, HLA-J, HLA-K, HLA-L, HLA-V, HLA-DRB2, HLA-DRB3, HLA-DRB4, HLA-DRB5, HLA-DRB6, HLA-DRB7, HLA-DRB8, HLA-DRB9, HLA-DOA, HLA-DOB, HLA-DMA, HLA-DMB, MICA, MICB, TAP1*, and *TAP2*), along with high-density SNP data in the MHC region by genotyped using the Illumina HumanCoreExome BeadChip (v1.1; Illumina) of 1,120 unrelated individuals of Japanese ancestry.^3^ It was phased using Beagle imputation software. We excluded 2 individuals’ data of which sides of some HLA alleles were inconsistent among different resolutions.

We used 908 individuals of Japanese ancestry with 4-digit resolution alleles of classical HLA genes (*HLA-A, HLA-B, HLA-C, HLA-DRB1, HLA-DQA1, HLA-DQB1, HLA-DPA1*) based on SSO method to benchmark the imputation performance when the Japanese panel was applied to an independent dataset. The dataset was used as an HLA reference panel in our previous study.^6^ It contains high-density SNP data genotyped using four SNP genotyping arrays (the Illumina HumanOmniExpress BeadChip, the Illumina HumanExome BeadChip, the Illumina Immunochip, and the Illumina HumanHap550v3 Genotyping BeadChip). It was distributed in a phased condition with Beagle format. Samples with missing genotype data for a locus were excluded in the accuracy evaluation of the locus. This study was approved by the ethics committee of Osaka University Graduate School of Medicine with written informed consent obtained from all participants.

#### (ii) The Type 1 Diabetes Genetics Consortium (T1DGC) reference panel

The T1DGC panel contains 5,868 SNPs (genotyped using Illumina Immunochip) and 4-digit resolution HLA typing data of classical HLA genes (*HLA-A, HLA-B*, and *HLA-C* for class I; *HLA-DPA1, HLA-DPB1, HLA-DQA1, HLA-DQB1*, and *HLA-DRB1* for class II) based on SSO method of 5,225 unrelated individuals of European ancestry.^22^ It was distributed in a phased condition with Beagle format. We excluded 103 individuals’ data of which sides of some HLA alleles were inconsistent among different resolutions.

### HLA imputation in 1000 Genomes Project data

We used Phase III 1000 Genomes Project (1KGv3) cohort as independent data to evaluate imputation accuracy. It comprises 2,554 individuals of 5 different super populations (AFR, AMR, EAS, EUR, and SAS). We obtained NGS-based 4-digit resolution HLA typing data for classical HLA genes (*HLA-A, HLA-B*, and *HLA-C* for class I; *HLA-DRB1* and *HLA-DQB1* for class II). The process of HLA typing has been described elsewhere.^59^ We evaluated imputation accuracy for individual populations based on their allele frequencies. Samples containing ambiguous alleles for a locus were excluded in the accuracy evaluation of that locus.

We experimentally constructed a mixed panel by merging the Japanese and T1DGC panels to assess imputation accuracy in diverse populations of 1KGv3. Considering the disparity in allele frequency of SNPs between two populations, we removed all palindromic SNVs to align the strands correctly when merging reference panels. We used 1,445 SNPs for imputation which were consistent with 1KGv3 genotype data. We used the same 1,445 SNPs for imputation to compare the accuracies in the same condition when we evaluated imputation accuracy using the Japanese panel.

### T1D GWAS data in the Japanese population

The BioBank Japan (BBJ) is a multi-institutional hospital-based registry that comprises DNA, serum, and clinical information of approximately 200,000 individuals of Japanese ancestry recorded from 2003 to 2007.^60,61^ We used GWAS data from 831 cases who had record of T1D diagnosis and 61,556 controls of Japanese genetic ancestry enrolled in the BBJ Project. The controls were same as those enrolled in our previous study that investigated the association of the MHC region with comprehensive phenotypes, and the number of T1D cases was increased.^3^ The process of patient registration, the GWAS data, and the QC process are described elsewhere.^60–62^

### T1D GWAS data in the British population

The UK Biobank (UKB) comprises health-related information approximately 500,000 individuals aged between 40–69 recruited from across the United Kingdom from 2006 to 2010.^63^ We used GWAS data of 732 T1D patients and 353,727 controls of British genetic ancestry enrolled in UKB. We selected T1D patients as individuals who were diagnosed with insulin-dependent diabetes mellitus in hospital records, and eliminated individuals with non-insulin-independent diabetes mellitus in hospital records and type 2 diabetes in self-reported diagnosis. The controls were individuals with no record of any autoimmune diseases in hospital records or in self-reported diagnosis. We included only individuals of British ancestry according to self-identification and criteria based on principal component (PC).^64^ We excluded individuals of ambiguous sex (sex chromosome aneuploidy and inconsistency between self-reported and genetic sex), and outliers of heterozygosity or call rate of high quality markers.

### Imputation of the HLA variants of GWAS data of T1D cases and controls

In this study, we defined the HLA variants as SNVs in the MHC region, classical 2-digit and 4-digit biallelic HLA alleles, biallelic HLA amino acid polymorphisms corresponding to the respective residues, and multiallelic HLA amino acid polymorphisms for each amino acid position. We applied DEEP*HLA to the GWAS data to determine classical 2-digit and 4-digit biallelic HLA alleles. The dosages of biallelic HLA amino acid polymorphisms corresponding to the respective residues and multiallelic HLA amino acid polymorphisms of each amino acid position were determined from the imputed 4-digit classical allele dosages. We applied post-imputation filtering as the biallelic alleles in which *r*^*2*^ accuracy in 10-fold cross-validation was lower than 0.7. The SNVs in the MHC region were imputed using minimac3 (version 2.0.1) after pre-phased with Eagle (version 2.3). We applied stringent post-imputation QC filtering of the variants (minor allele frequency ≥ 0.5% and imputation score Rsq ≥ 0.7). For trans-ethnic fine-mapping, we integrated results of the imputation of individual cohorts by including the HLA genes, amino acid position, and SNVs that were typed in both reference panels. Regarding the HLA alleles and amino acid polymorphisms, those present in one population were regarded as absent in the other population. Considering the disparity in allele frequency of SNVs among different populations, we removed all palindromic SNVs to correctly align the strands.

### Association testing of the HLA variants

We assumed additive effects of the allele dosages on the log-odds scales for susceptibility to T1D; and evaluated associations of the HLA variants with the risk of T1D using a logistic regression model. To robustly account for potential population stratification, we included the top 10 PCs obtained from the GWAS genotype data of each cohort (not including the MHC region) as covariates in the regression model. We also included ascertainment centre and genotyping chip for UKB as covariates. For trans-ethnic analysis, PC terms for each other population were set to 0, and a categorical variable indicating a population was added as a covariate. We also included the sex of individuals as a covariate.

To evaluate independent risk among the HLA variants and genes, we conducted a forward-type stepwise conditional regression analysis that additionally included the associated variant genotypes as covariates. When conditioning on HLA gene(s), we included all the 4-digit alleles as covariates to robustly condition the associations attributable to the HLA genes, as previously described.^3,14^ When conditioning on the specific HLA amino acid position(s), we included the multiallelic variants of the amino acid residues. We applied a forward stepwise conditional analysis for the HLA variants and then HLA genes, based on a genome-wide association significance threshold (*P* = 5.0 × 10^−8^). A previous study reported that the T1D risk was strongly associated with a combination of variants in the region of *HLA-DRB1, -DQA1*, and *-DQB1*, where the variants have strong LD to each other.^12^ In such a situation, conditioning on all the 4-digit alleles of a single HLA gene might inadvertently blind the association of alleles of other HLA genes; therefore, we conditioned on a set of individual HLA variants rather than an each HLA gene when analyzing this region.

We tested a multivariate full regression model by including the risk-associated HLA variants in *HLA-DRB1, HLA-DQB1, HLA-A*, and *HLA-B*, which were identified through the stepwise regression analysis. We excluded the most frequent residue in the British cohort from each amino acid position as the reference allele when we included amino acid polymorphisms in the model. Phenotypic variance explained by the identified risk-associated HLA variants was estimated on the basis of a liability threshold model assuming a population-specific prevalence of T1D and using the effect sizes obtained from the multivariate regression model.

## URLs

DEEP*HLA, https://github.com/tatsuhikonaito/DEEP-HLA

Pytorch, http://pytorch.org/

SNP2HLA, http://software.broadinstitute.org/mpg/snp2hla/

HIBAG, https://www.bioconductor.org/packages/release/bioc/html/HIBAG.html

Eagle, https://data.broadinstitute.org/alkesgroup/Eagle/

Minimac3, https://genome.sph.umich.edu/wiki/Minimac3

BioBank Japan, https://biobankjp.org/english/index.html

UK Biobank, https://www.ukbiobank.ac.uk/

UCSF Chimera, https://www.cgl.ucsf.edu/chimera/

