## Supplementary Information for "A multi-task convolutional deep learning method for HLA allelic imputation and its application to trans-ethnic MHC fine-mapping of type 1 diabetes"

Naito T et al.

Corresponding to Yukinori Okada

### Supplementary Note 1. Extended evaluations of imputation accuracies of DEEP*HLA

To benchmark the accuracies of DEEP*HLA more comprehensively, we tested its performance in various aspects.

#### a. Effects of down-sampling on accuracy of DEEP*HLA

We evaluated the effects of down-sampling of training data on accuracies of DEEP*HLA. We performed 10-fold cross-validation for both reference panels and independent samples for our Japanese reference panel, where only part of an original training fold was used as a training fold. We tested a down-sampling rate of 90, 80, 70, 60, 50, 40, 30, 20, and 10%. The results are shown in **Supplementary Fig. 1**.

#### b. Comparison with single-task neural networks, and multi-task neural networks with shuffled groupings

We evaluated the advantages of the multi-task learning with grouping. The multi-task learning would be effective mainly in our Japanese reference panel in which more HLA gene loci were genotyped than T1DGC panel; thus, we tested only for our Japanese panel.

##### b.1. Comparison with single-task neural networks

We tested the performance of single-task neural networks that imputed all genes separately. To perform a fair comparison, the input regions were set to the same as DEEP*HLA with the original grouping. As shown in **Supplementary Fig. 11a**, all the accuracies were lower than the multi-task DEEP*HLA in all the ranges of allele frequencies. Moreover, the mean training time in the cross-validation was 192 min per one iteration, which was over 5 times longer than the multi-task learning (36 min).

**b.2. Comparison with multi-task neural networks with shuffled grouping**

To evaluate the advantage of the original grouping, we evaluated the performance of models with shuffled grouping. We investigated two cases: (A) shuffling HLA genes between group 1 and 2, and between 3 and 4; (B) shuffling HLA genes among group 1, 2, 3, and 4. We tested 5 different groupings for each case. As shown in **Supplementary Fig. 11b**, DEEP*HLA with the original grouping was significantly outperformed those with the shuffled groupings. The groupings (A) tended to perform better than the groupings (B). These results suggest the importance of grouping based on the physical distance and LD structures.

**c. Comparison among different input window sizes**

We benchmarked DEEP*HLA with different window sizes of 250, 750, and 1,000 kb in addition to 500 kb (**Supplementary Fig. 12**). Although the optimal window size might vary by locus in rare allele, there was no significant difference overall.

**d. Strict cross-validation including haplotype pre-phasing**

HLA references panels in a phased condition with all the subjects were used for the cross-validation shown in the main text. In a real scenario, however, reference data (i.e. a training fold) and target data (i.e. a validation fold) are more likely to be independently phased. Thus, we conducted stricter cross-validation for the accuracy of DEEP*HLA in which each training data was pre-phasing after separation. As shown in **Supplementary Fig. 15**, there were no significant overall changes in the accuracies but a slight decline in alleles with a frequency < 0.5%. Especially, alleles with a frequency < 0.1% correspond to doubleton (or singleton) in the Japanese panel; thus, separate pre-phasing is likely to have a slight effect on imputation performance.

**Supplementary Note 2. An illustration of accuracy metrics for imputed dosages used in our study**

Based on a cross-tabulation table (**Supplementary Fig. 14a**), we defined a per-allele sensitivity of imputed dosage as

$$Se(A) = \frac{\sum_{j=1}^{m} D_j(A)}{m_A} = \frac{TP}{TP + FN}$$

where *m* denotes the number of true observations of allele *A* in total sample, and $D_i$ represents imputed dosage of allele *A* in individual haplotype *j* which has allele *A*. *TP* (true positive) and *FN* (false negative) are illustrated in the cross-tabulation table.

Accuracy of a locus defined in the paper of SNP2HLA *Acc* is calculated by summing across all individuals the dosage of each true allele in the individual (i.e. the sum of true positives of all individual alleles), divided by the total number of observations. As shown in **Supplementary Fig**. **14b**, it is consistent with a weighted-mean of the per-allele sensitivity by allele frequencies as

$$\begin{aligned} Acc &= \frac{\sum_{i=1}^{n}\left(D_i(A1_{i,L}) + D_i(A2_{i,L})\right)}{2n} \\ &= \frac{\sum_a TP_a}{2n} \\ &= \sum_a \left(\frac{TP_a + FN_a}{2n}\right)\left(\frac{TP_a}{TP_a + FN_a}\right) \\ &= \sum_a freq_a \cdot Se(a) \end{aligned}$$

where *n* denotes the number of individuals, $D_i$ represents the imputed dosage of an allele in individual *i*, and alleles $A1_{i,\,L}$ and $A2_{i,\,L}$ represent the true HLA alleles for individual *i* at locus *L*. $TP_\mathrm{a}$, $FN_\mathrm{a}$, and $freq_a$ denotes the true positive, false negative, and allele frequency of an allele *a*.  This is why we termed *Acc* as a sensitivity for each locus.

### Supplementary Figure 1. Evaluation of accuracy of DEEP*HLA in a down-sampling approach.

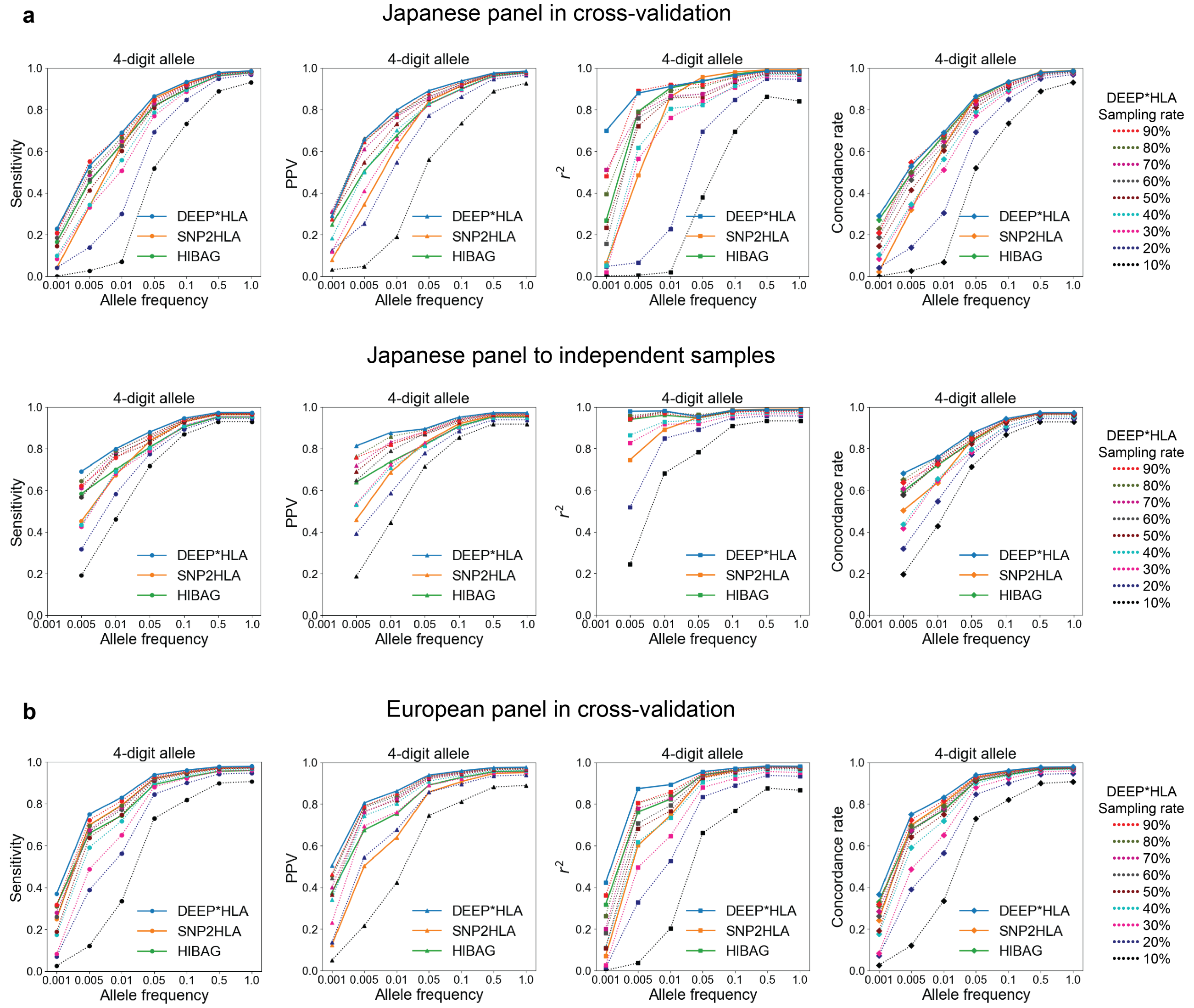

Accuracies evaluated with a down-sampling approach for the 4-digit allele in our Japanese reference panel (**a**) and T1DGC reference panel (**b**). Each colored dotted line represents the accuracies of DEEP*HLA with a certain sampling rate as shown in the right legend. For each metric, those for alleles of which frequency is less than a value on the horizontal axis are shown on the vertical axis.

**Supplementary Figure 2. Accuracy evaluation of HLA imputation methods in 1000 Genome Projects data.**

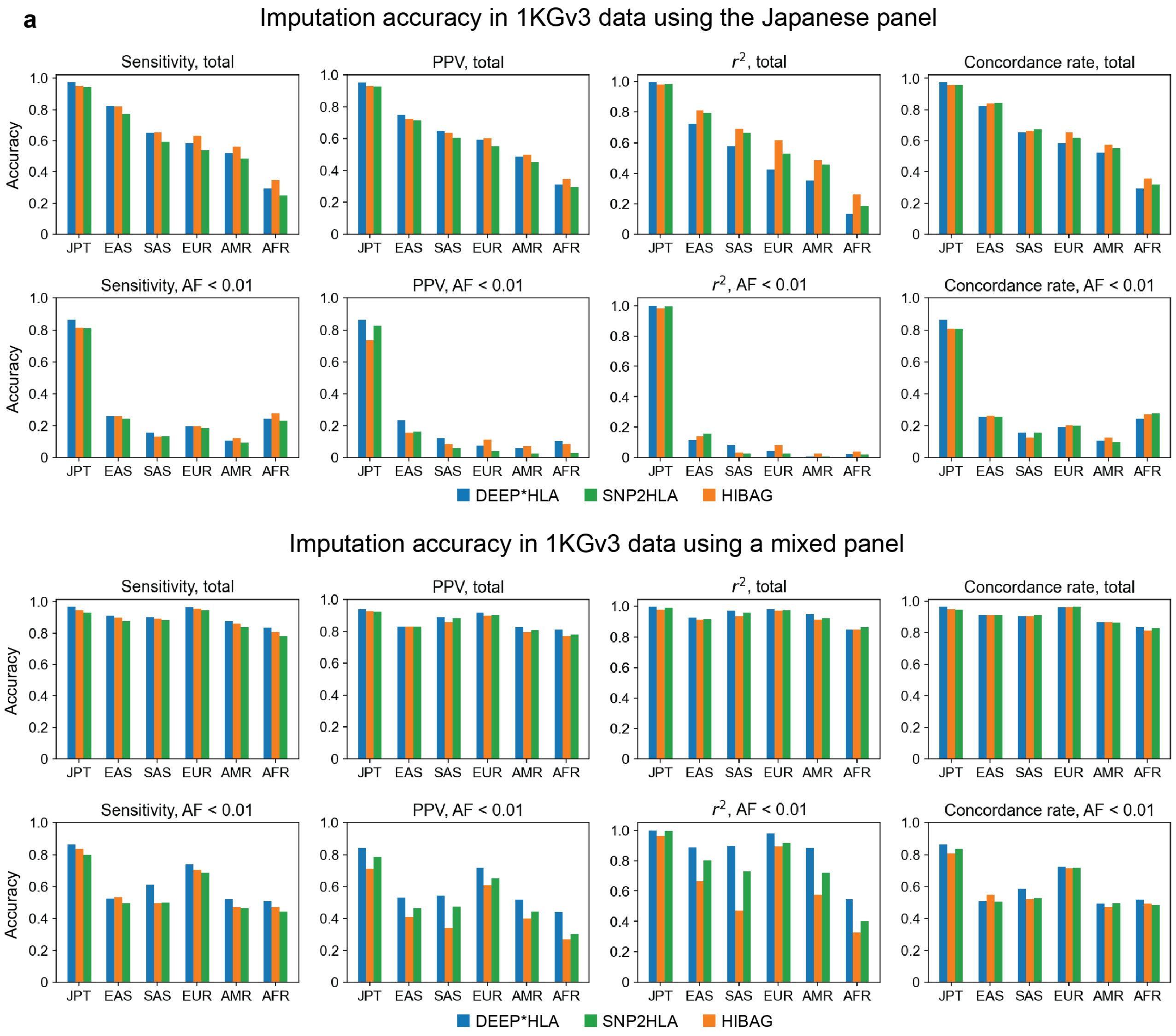

Each bar plot represents the imputation accuracy in diverse populations using our Japanese reference panel (a), and a mixed panel of the Japanese and European panels (b), which were averaged in all alleles (upper) and alleles with a frequency < 1% (lower). EAS represents the EAS cohort excluding the JPT cohort.

**Supplementary Figure 3. Receiver operating characteristic curves for ability for entropy-based uncertainty and genotype dosage of discriminating incorrectly imputed 4-digit alleles.**

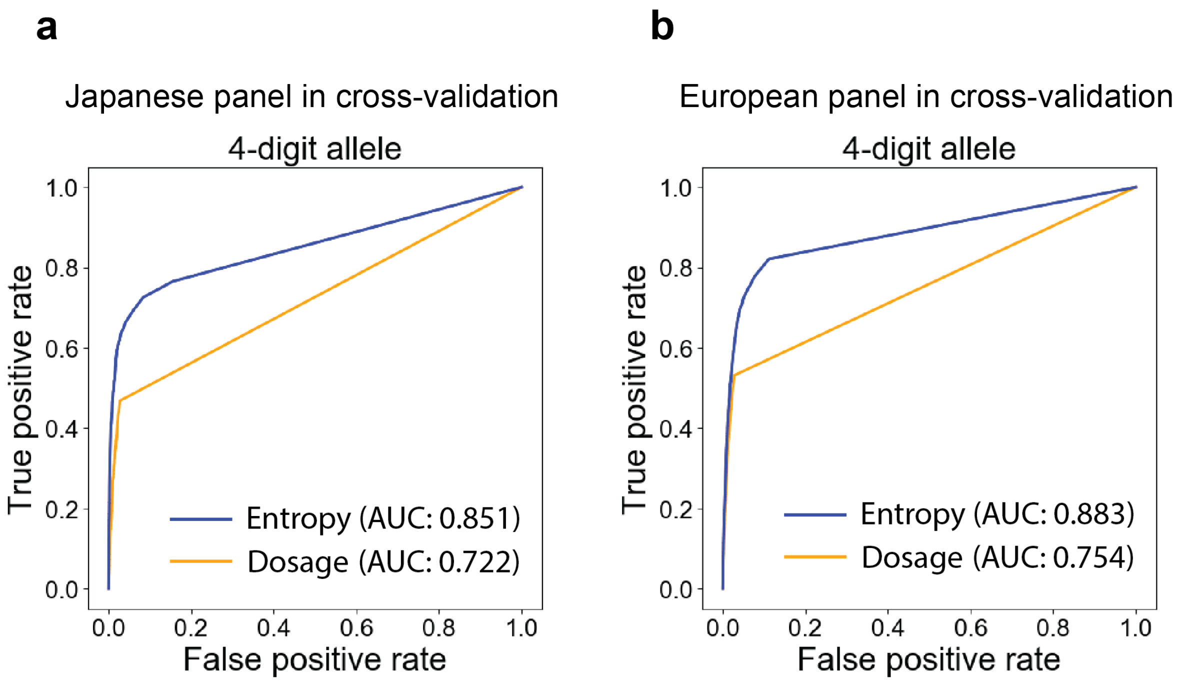

The entropy-based uncertainty was able to discriminate incorrectly imputed 4-digit alleles with a higher accuracy than genotype dosage both in the Japanese panel (**a**) and T1DGC panel (**b**).

**Supplementary Figure 4. T1D risk-associated variants in HLA-DRB1, -DQA1, and -DQB1 identified by stepwise conditional association analysis.**

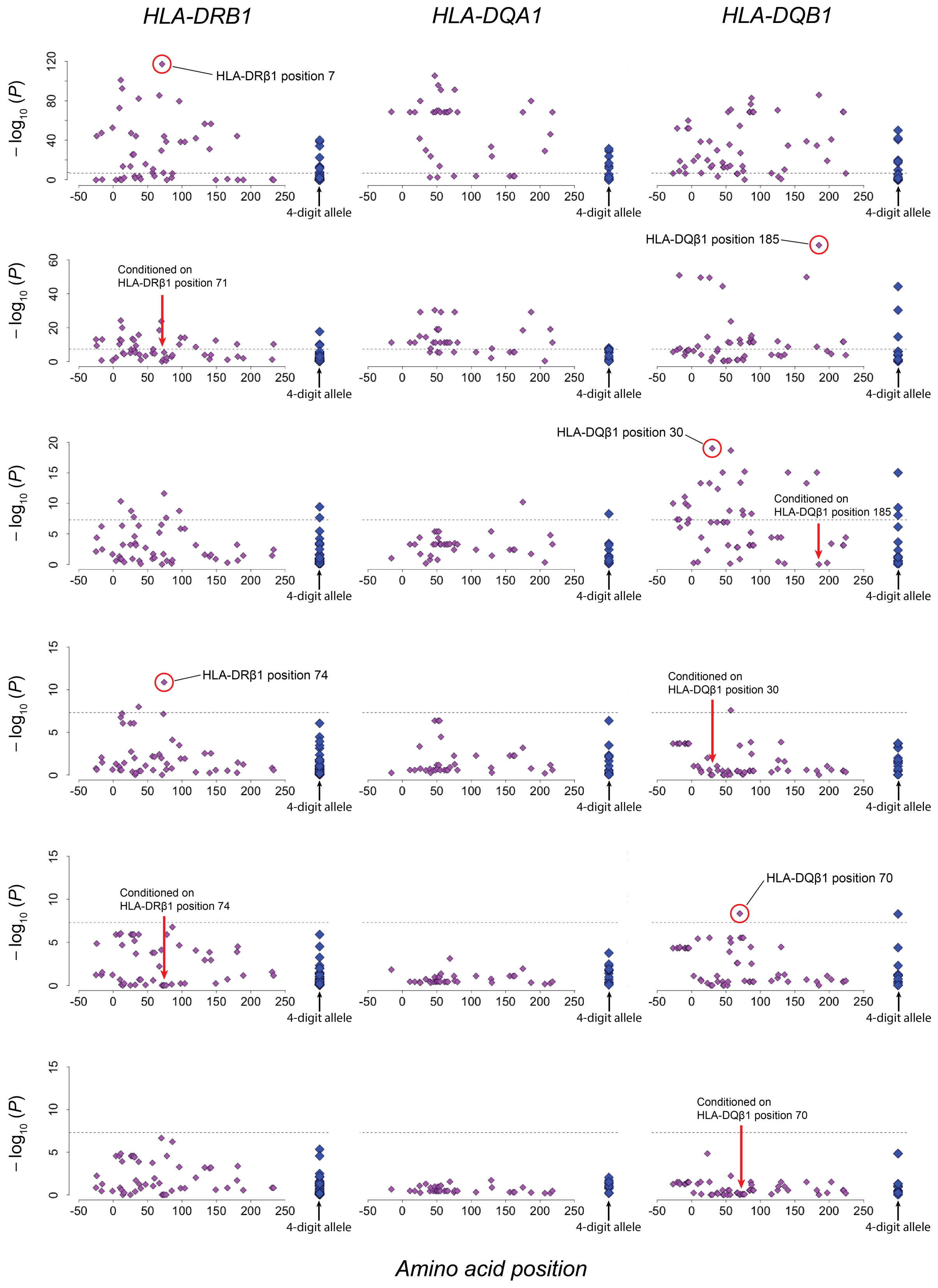

Diamonds represent the $-\log_{10}$ ($P$) values of the amino acid polymorphisms (purple) and classical alleles (blue) for the tested HLA gene. For amino acid polymorphisms, the smallest $P$ values among the binary $P$ values (= $P_{binary}$) and omnibus $P$ values (= $P_{omnibus}$) at each position are indicated. An allele of the smallest P values at each step is displayed in red circle. The dashed horizontal lines represent the genome-wide significance threshold of $P = 5.0\times10^{-8}$.

**Supplementary Figure 5. Comparison of odds ratios of T1D risk-associated variants in *HLA-DRB1* and *-DQB1* between Japanese and Europeans.**

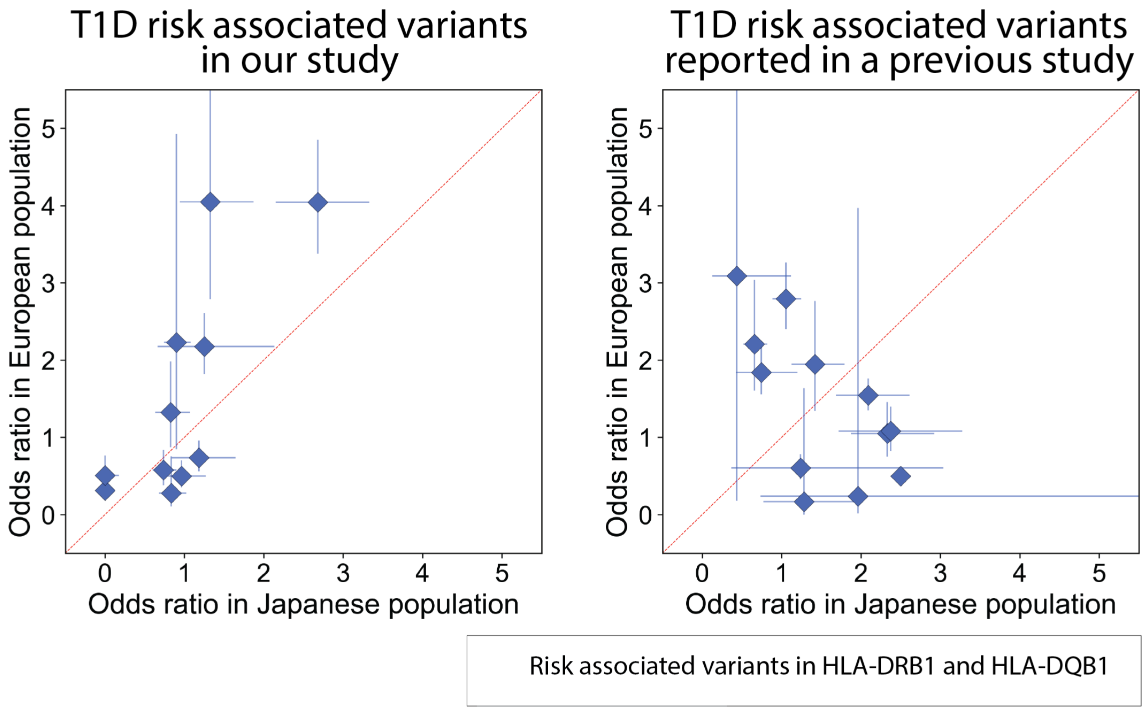

Odds ratios (ORs) of the variant observed in our study (left) and of reported previously (left) in HLA-DRB1 and HLA-DQB1 are plotted based on those in Japanese (horizontal axis) and Europeans (vertical axis). For each variant, its 95% confidence intervals of ORs are plotted as error bars.

**Supplementary Figure 6. An association plots of HLA variants with T1D in the MHC region for the BBJ cohort.**

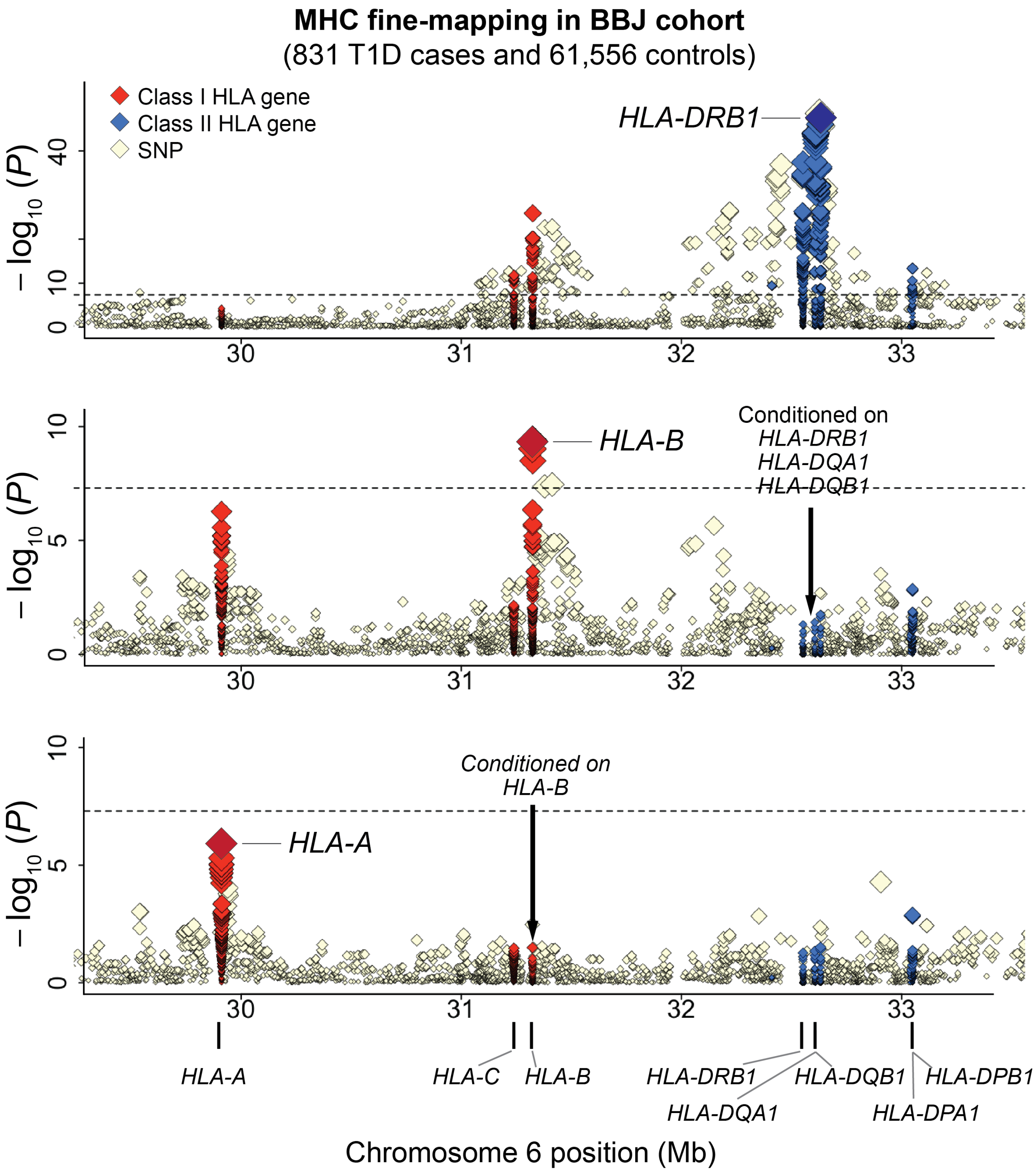

**Supplementary Figure 7. An association plots of HLA variants with T1D in the MHC region for the UKB cohort.**

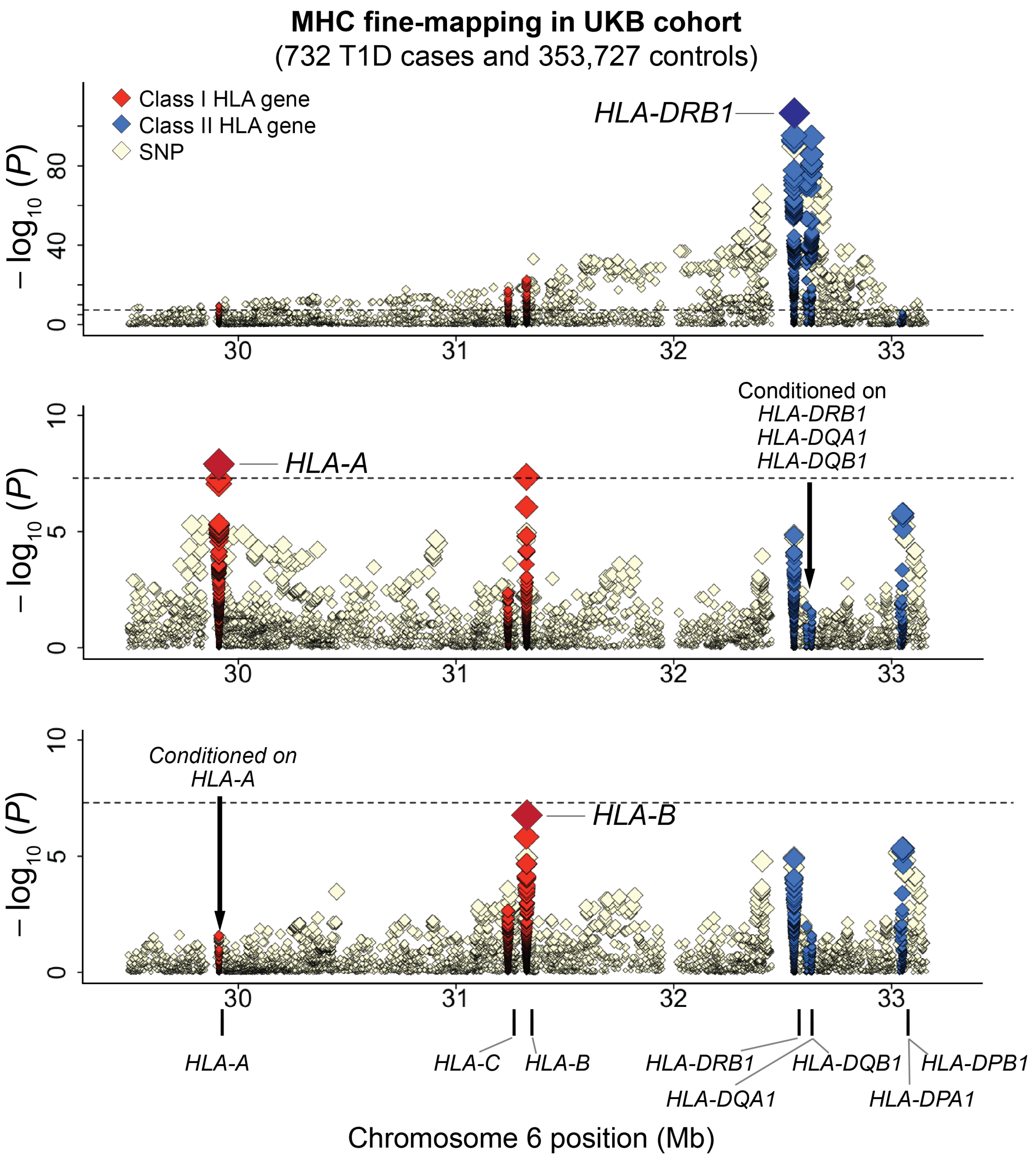

**Supplementary Figure 8. T1D risk-associated variants in *HLA-DRB1*, *-DQA1*, and *-DQB1* identified by stepwise conditional association analysis in BBJ cohort.**

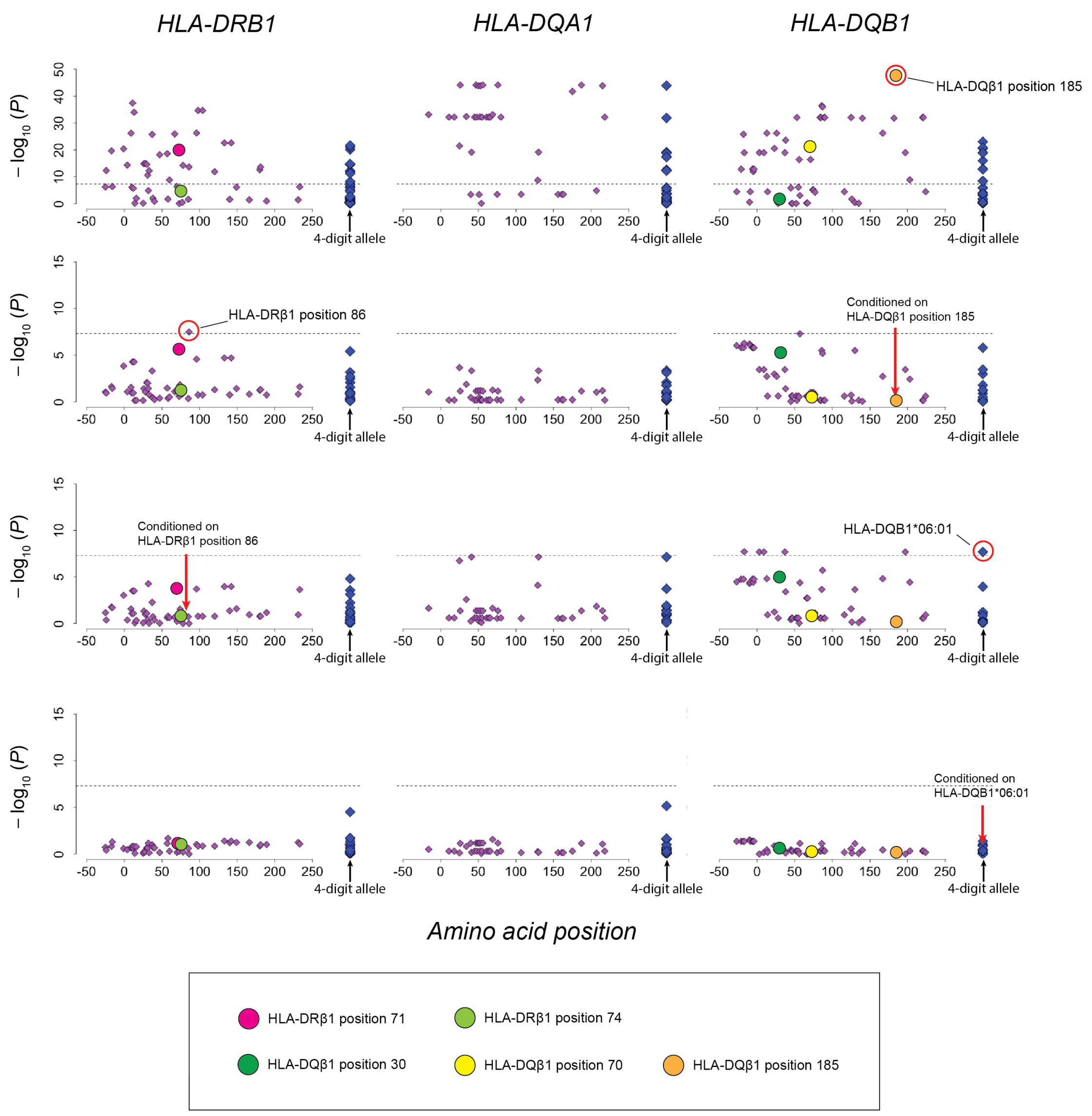

**Supplementary Figure 9. T1D risk-associated variants in *HLA-DRB1*, *-DQA1*, and *-DQB1* identified by stepwise conditional association analysis in UKB cohort.**

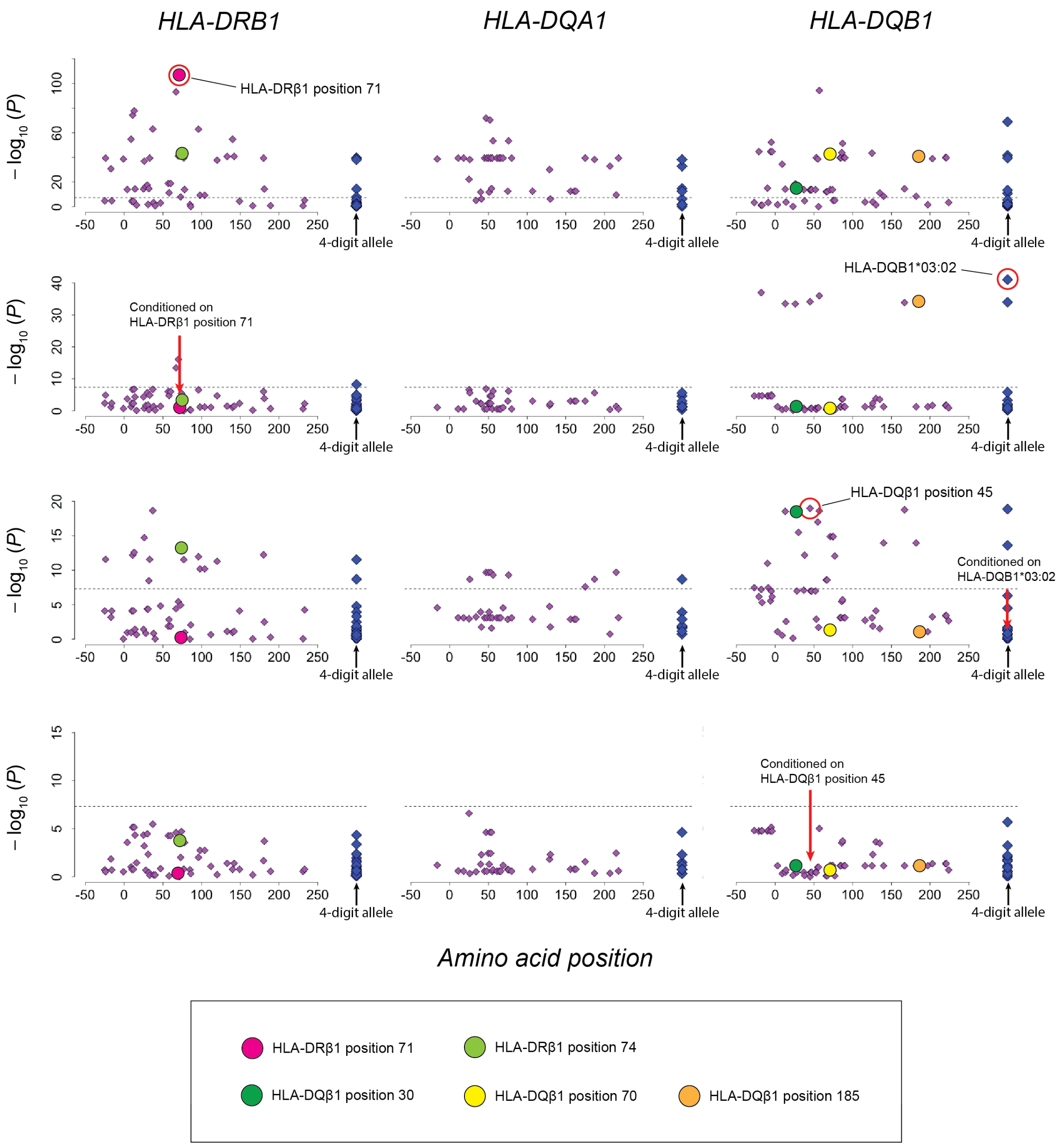

**Supplementary Figure 10. Three-dimensional illustration of T1D risk-associated amino acid positions identified by trans-ethnic MHC fine-mapping.**

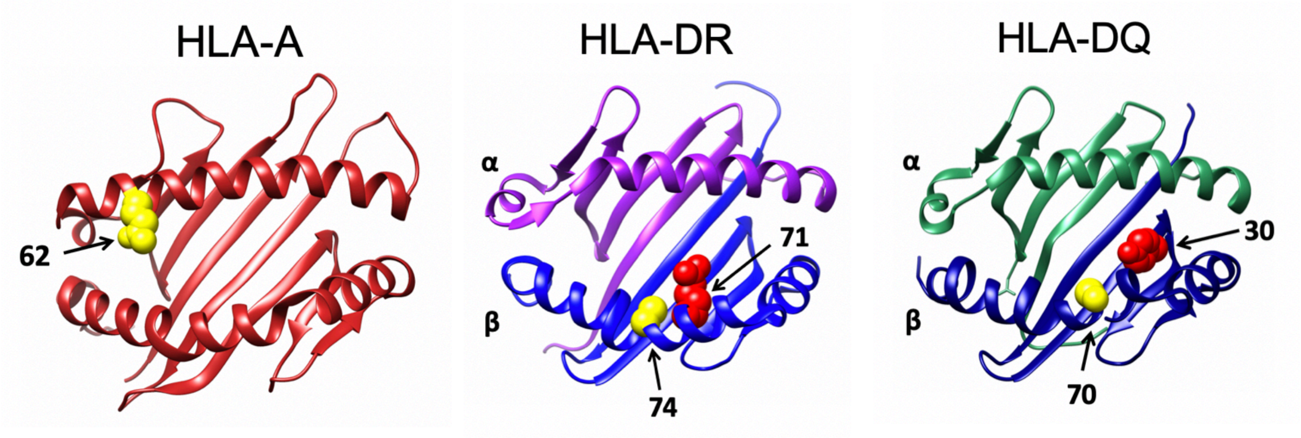

The protein structures of HLA-A, HLA-DR, and HLA-DQ are based on Protein Data Bank entries 2BVP, 3PDO, and 1UVQ, respectively, which were displayed using UCSF Chimera version 1.14. Residues at the T1D risk-associated amino acid positions are colored yellow or red (arrows).

**Supplementary Figure 11. Comparison of DEEP*HLA of the original grouping with single-task neural networks and those of shuffled groupings.**

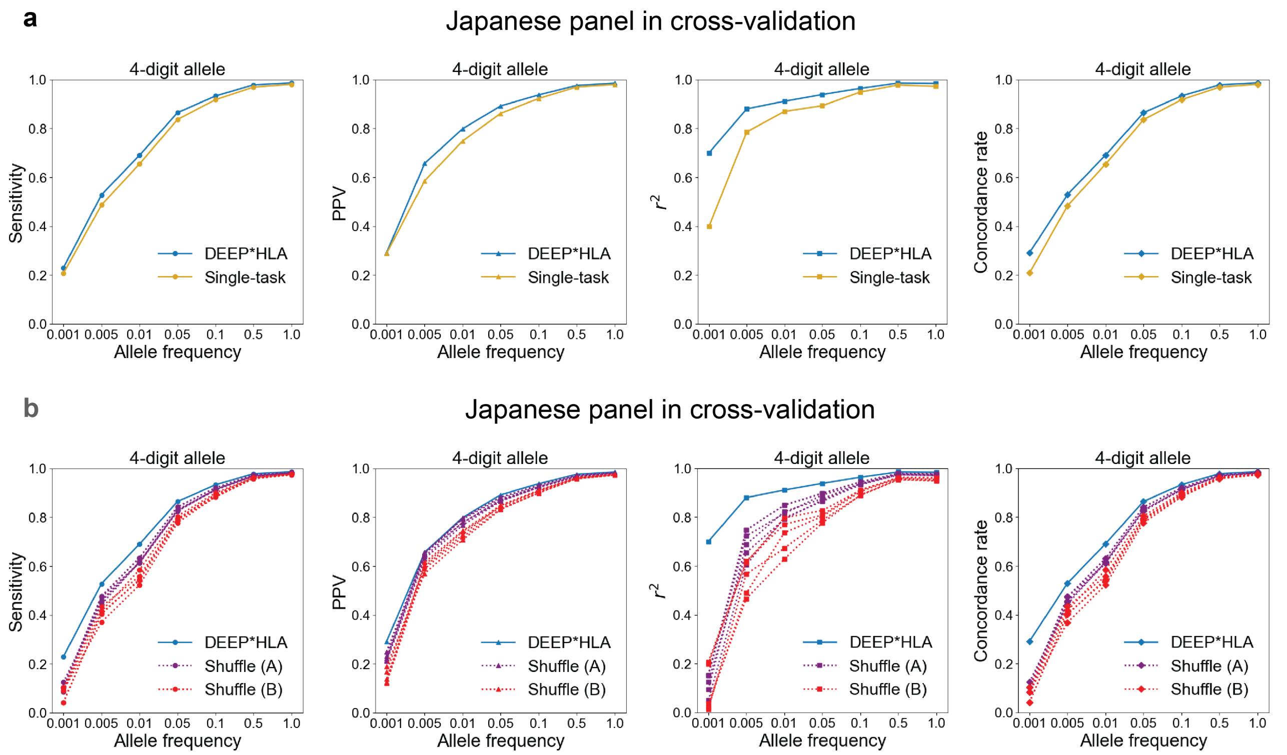

Comparison of DEEP*HLA of the original grouping with single-task neural networks (a) and those of shuffled groupings (**b**) in accuracies of for the 4-digit alleles evaluated in the Japanese panel in cross-validation. For each metric, those for alleles of which frequency is less than a value on the horizontal axis are shown on the vertical axis.

**Supplementary Figure 12. Comparison of DEEP*HLA with different input ranges.**

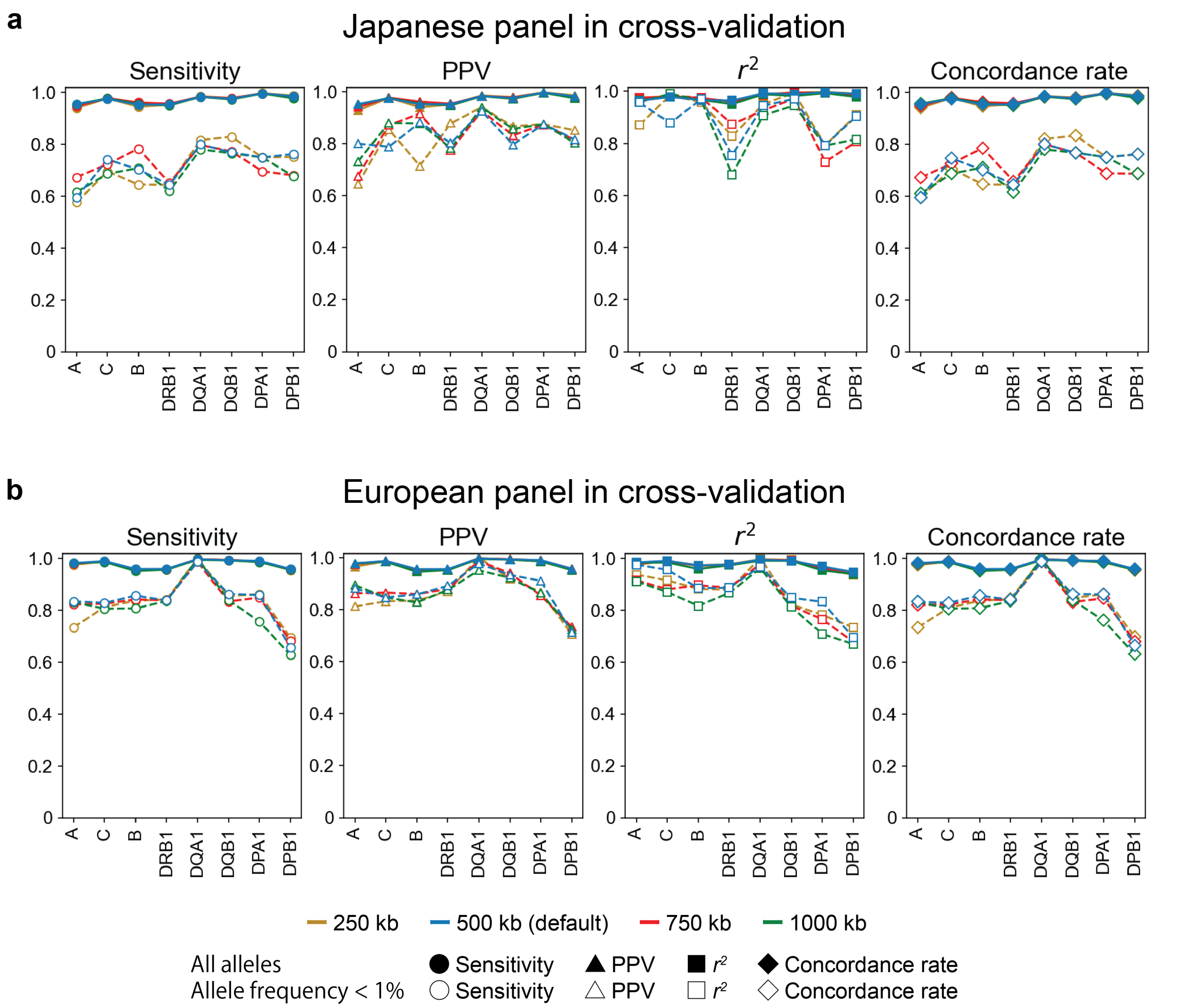

Each panel represents the accuracies of DEEP*HLA with different input ranges in 8 classical HLA genes evaluated in the Japanese panel in cross-validation (a) and the European panel in cross-validation (b). Solid and dashed lines correspond to the accuracies of all the allele and allele frequency < 1%, respectively.

**Supplementary Figure 13. Data separation for training DEEP*HLA in a 10-fold cross-validation.**

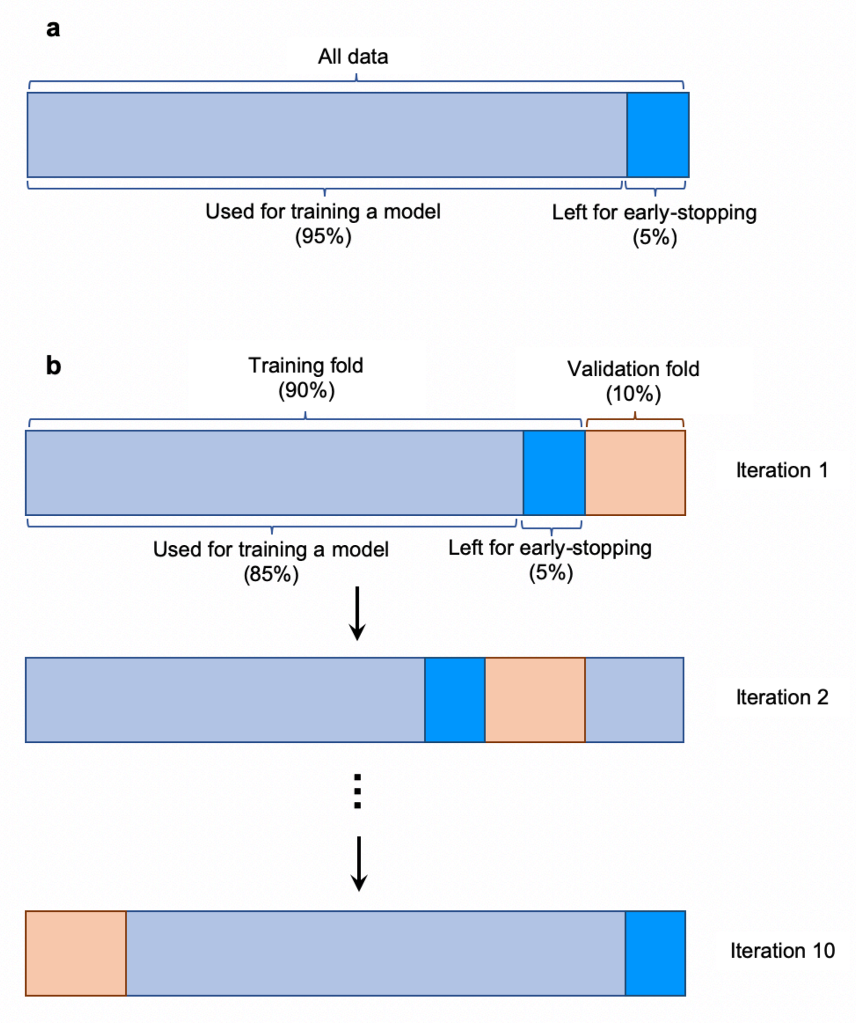

(a) In a general way of training a DEEP*HLA model, 5% of data are left as sub-validation data for early-stopping training (b) In 10-fold cross-validation, sub-validation data were separated from the training fold for robust accuracy evaluation. Thus, 85% of whole data were used for training a model.

**Supplementary Figure 14. An illustration of accuracy metrics for imputed dosages.**

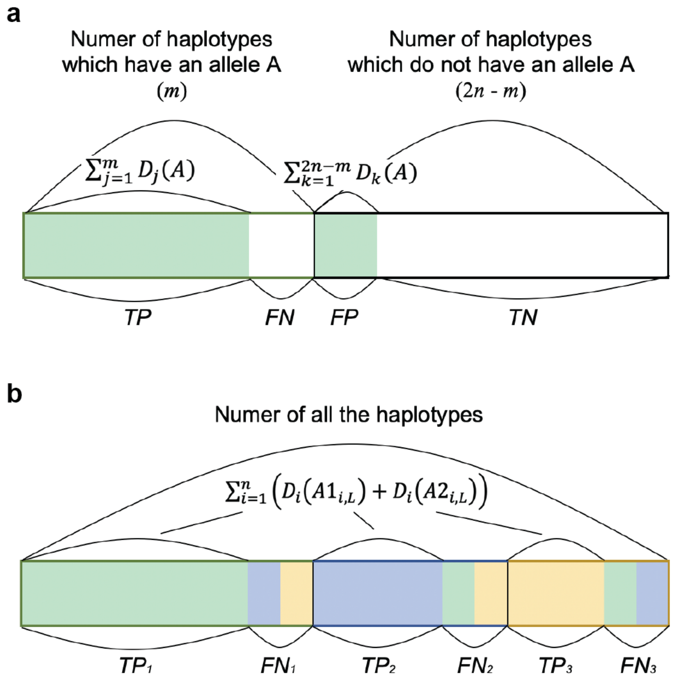

| | True A | True non-A | |
|---|---|---|---|
| Imputed as A | *TP* | *FP* | *FP + TN* |
| Imputed as non-A | *FN* | *TN* | *FN + TN* |
| | *TP + FN* | *FP + TN* | 2$n$ |

A box represents a sum of allelic dosage of all the haplotypes with colored as imputed dosages of an allele in biallelic (**a**) and multi-allelic representation of 3 alleles (**b**). The frame color represents true observation of each allele. We defined accuracies of sensitivity and PPV based on a cross-tabulation table (**a**, right). TP, true positive; FP, false positive; FN, false negative; TN, true negative.

**Supplementary Figure 15. Imputation accuracy of DEEP*HLA in strict cross-validation.**

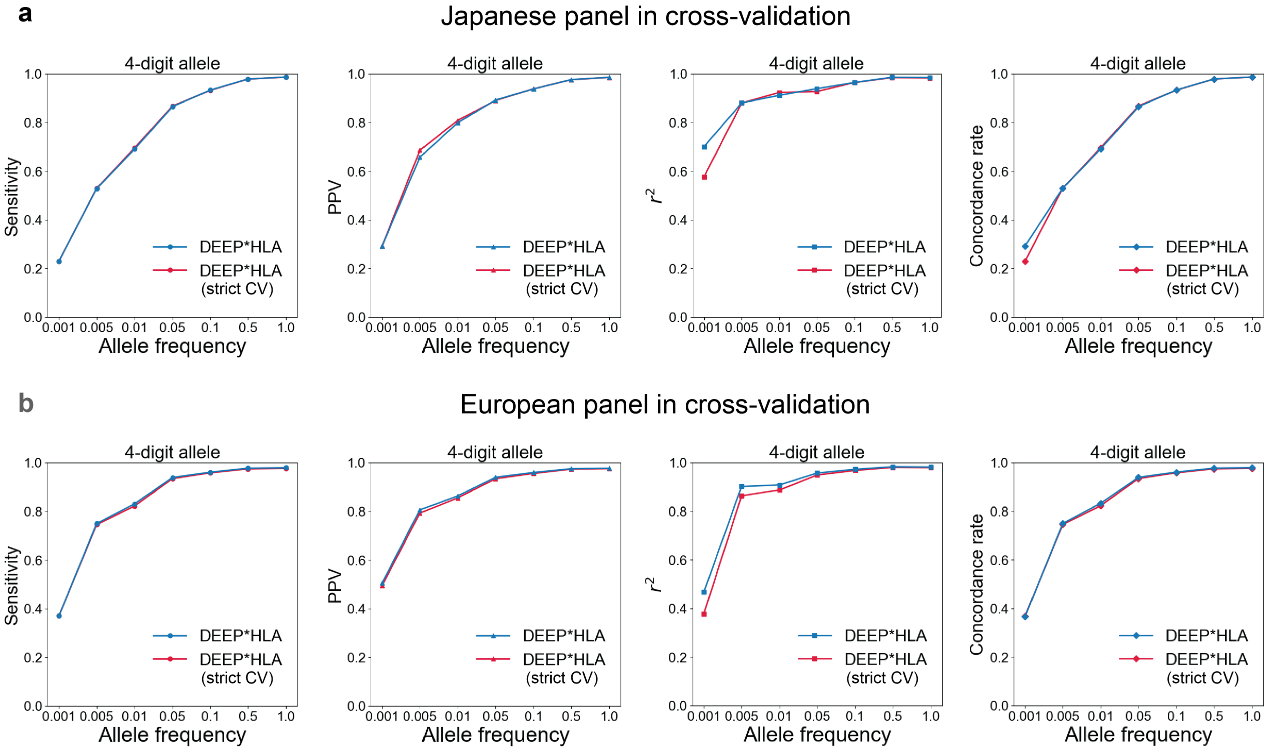

Comparison in accuracy of DEEP*HLA between cross-validation with and without considering pre-phasing process in the Japanese panel (**a**) and the European panel (**b**). For each metric, those for alleles of which frequency is less than a value on the horizontal axis are shown on the vertical axis. CV, cross-validation.

**Supplementary Table 1. Summary for measurement of processing time and maximum memory usages of HLA imputation methods**

| n | Phasing GWAS data time (min) | Imputation time (min) | | | Total processing time (min) | | | Maximum memory usage (MB) | | |
|---|---|---|---|---|---|---|---|---|---|---|
| | | DEEP*HLA | SNP2HLA | HIBAG | DEEP*HLA | SNP2HLA | HIBAG | DEEP*HLA | SNP2HLA | HIBAG |
| **1,000** | 4.9 | 1.6 | 12.0 | 12.4 | 159.8 | 12.0 | 3285.0 | 3849.6 | 10877.9 | 162.7 |
| **2,000** | 9.8 | 3.0 | 32.5 | 24.9 | 166.2 | 32.5 | 3297.5 | 3946.8 | 14117.7 | 245.8 |
| **5,000** | 33.9 | 7.5 | 124.3 | 62.3 | 194.7 | 124.3 | 3334.9 | 4135.4 | 18133.1 | 466.2 |
| **10,000** | 72.5 | 15.5 | 481.7 | 123.6 | 241.3 | 481.7 | 3396.2 | 4437.4 | 37697.2 | 833.7 |
| **20,000** | 154.6 | 34.3 | 2261.5 | 250.5 | 342.2 | 2261.5 | 3523.1 | 5105.6 | 80094.4 | 1419.8 |
| **50,000** | 408.4 | 78.1 | NA | 620.0 | 639.8 | NA | 3892.6 | 7247.0 | NA | 1724.5 |
| **100,000** | 869.5 | 166.9 | NA | 1253.6 | 1189.8 | NA | 4526.2 | 12442.8 | NA | 3502.9 |

In DEEP*HLA, the total processing times were determined by summing phasing GWAS data time (by Eagle), training time (153 min), and imputation time. In HIBAG, the sums of training time (3,273 min) and imputation were regarded as the total processing time.

### Supplementary Table 2. Results of linear regression analysis for association of AUC for distant-dependent LD decay with imputation accuracy metrics

| | | | Sensitivity | | PPV | | $r^2$ | | Concordance rate | |
|---|---|---|---|---|---|---|---|---|---|---|
| | | | Beta | P | Beta | P | Beta | P | Beta | P |
| Japanese | DEEP*HLA | AUC for LD decay | 0.140 | $1.8 \times 10^{-10}$ | 0.108 | $5.7 \times 10^{-9}$ | 0.124 | $6.6 \times 10^{-10}$ | 0.137 | 2.1 × 10-$^{10}$ |
| | | Allele frequency | 0.083 | $7.6 \times 10^{-5}$ | 0.051 | 0.0049 | 0.062 | 0.0014 | 0.083 | $7.9 \times 10^{-5}$ |
| | SNP2HLA | AUC for LD decay | 0.190 | $3.8 \times 10^{-18}$ | 0.180 | $1.7 \times 10^{-16}$ | 0.180 | $2.3 \times 10^{-16}$ | 0.200 | $2.1 \times 10^{-18}$ |
| | | Allele frequency | 0.095 | $7.5 \times 10^{-6}$ | 0.097 | $2.6 \times 10^{-6}$ | 0.086 | $5.2 \times 10^{-5}$ | 0.094 | $1.9 \times 10^{-5}$ |
| | DEEP*HLA | AUC for LD decay† | 0.150 | $5.9 \times 10^{-11}$ | 0.105 | $1.5 \times 10^{-7}$ | 0.119 | $3.0 \times 10^{-8}$ | 0.146 | $6.5 \times 10^{-11}$ |
| | | Allele frequency | 0.066 | 0.0022 | 0.043 | 0.024 | 0.053 | 0.0090 | 0.066 | 0.0023 |
| | SNP2HLA | AUC for LD decay† | 0.190 | $6.2 \times 10^{-16}$ | 0.180 | $1.0 \times 10^{-15}$ | 0.190 | $1.2 \times 10^{-15}$ | 0.200 | $5.9 \times 10^{-16}$ |
| | | Allele frequency | 0.078 | $5.0 \times 10^{-4}$ | 0.079 | $2.6 \times 10^{-4}$ | 0.066 | 0.0026 | 0.076 | $9.8 \times 10^{-4}$ |
| | DEEP*HLA | LD max within 100 SNPs | 0.211 | $1.0 \times 10^{-21}$ | 0.140 | $2.3 \times 10^{-12}$ | 0.198 | $1.5 \times 10^{-21}$ | 0.211 | $1.3 \times 10^{-21}$ |
| | | Allele frequency | 0.026 | 0.19 | 0.021 | 0.26 | 0.011 | 0.54 | 0.026 | 0.19 |
| | SNP2HLA | LD max within 100 SNPs | 0.260 | $3.1 \times 10^{-29}$ | 0.250 | $6.8 \times 10^{-29}$ | 0.260 | $4.8 \times 10^{-31}$ | 0.270 | $2.0 \times 10^{-28}$ |
| | | Allele frequency | 0.034 | 0.099 | 0.036 | 0.064 | 0.019 | 0.33 | 0.032 | 0.13 |
| T1DGC | DEEP*HLA | AUC for LD decay | 0.130 | $5.4 \times 10^{-9}$ | 0.127 | $6.9 \times 10^{-9}$ | 0.127 | $1.2 \times 10^{-8}$ | 0.134 | $4.5 \times 10^{-9}$ |
| | | Allele frequency | 0.059 | 0.0080 | 0.043 | 0.044 | 0.065 | 0.0027 | 0.059 | 0.0083 |
| | SNP2HLA | AUC for LD decay | 0.170 | $4.2 \times 10^{-13}$ | 0.190 | $1.3 \times 10^{-19}$ | 0.180 | $1.4 \times 10^{-16}$ | 0.170 | $7.2 \times 10^{-13}$ |
| | | Allele frequency | 0.065 | 0.0040 | 0.087 | $1.1 \times 10^{-5}$ | 0.082 | $1.2 \times 10^{-4}$ | 0.065 | 0.0055 |
| | DEEP*HLA | AUC for LD decay† | 0.150 | $8.8 \times 10^{-12}$ | 0.140 | $1.2 \times 10^{-10}$ | 0.144 | $5.0 \times 10^{-11}$ | 0.154 | $8.1 \times 10^{-12}$ |
| | | Allele frequency | 0.053 | 0.014 | 0.040 | 0.058 | 0.060 | 0.0046 | 0.053 | 0.015 |
| | SNP2HLA | AUC for LD decay† | 0.180 | $1.5 \times 10^{-14}$ | 0.190 | $4.3 \times 10^{-19}$ | 0.180 | $8.7 \times 10^{-17}$ | 0.180 | $3.3 \times 10^{-14}$ |
| | | Allele frequency | 0.063 | 0.0042 | 0.090 | $5.9 \times 10^{-6}$ | 0.083 | $7.9 \times 10^{-5}$ | 0.064 | 0.0057 |
| | DEEP*HLA | LD max within 100 SNPs | 0.186 | $4.3 \times 10^{-14}$ | 0.154 | $1.6 \times 10^{-10}$ | 0.195 | $3.6 \times 10^{-16}$ | 0.187 | $4.7 \times 10^{-14}$ |
| | | Allele frequency | 0.006 | 0.78 | 0.006 | 0.80 | 0.006 | 0.79 | 0.006 | 0.78 |
| | SNP2HLA | LD max within 100 SNPs | 0.220 | $5.5 \times 10^{-18}$ | 0.250 | $5.3 \times 10^{-29}$ | 0.250 | $2.3 \times 10^{-25}$ | 0.220 | $5.0 \times 10^{-17}$ |
| | | Allele frequency | 0.008 | 0.74 | 0.021 | 0.30 | 0.015 | 0.50 | 0.008 | 0.74 |

† AUC of which window range equals the input region of DEEP*HLA
SE, standard error; AUC, area under the curve; LD, linkage disequilibrium.

### Supplementary Table 5. Associations of the previously reported HLA variants of *HLA-DRB1* and *HLA-DQB1* with T1D risk in trans-ethnic cohorts

| | Frequency (BBJ) | | Frequency (UKB) | | | | | |
|---|---|---|---|---|---|---|---|---|
| | Case | Control | Case | Control | OR (95% CI) | OR (95% CI) | P† | |
| HLA variant | n = 831 | n = 61,556 | n = 732 | n = 353,727 | BBJ | UKB | BBJ | UKB |
| HLA-DQB1 amino acid position 57 | | | | | | | | |
| Alanine | 0.13 | 0.10 | 0.61 | 0.36 | 1.05 (0.89-1.24) | 2.82 (2.43-3.29) | 0.52 | $8.2 \times 10^{-42}$ |
| Aspartic acid | 0.74 | 0.75 | 0.24 | 0.48 | (reference) | | | |
| Serine | 0.015 | 0.023 | 0.0014 | 0.0085 | 1.28 (0.78-2.02) | 0.17 (0.01-1.64) | 0.30 | 0.21 |
| Valine | 0.12 | 0.13 | 0.15 | 0.16 | 0.66 (0.53-0.81) | 2.21 (1.61-3.03) | $1.1 \times 10^{-4}$ | $8.2 \times 10^{-7}$ |
| HLA-DRB1 amino acid position 13 | | | | | | | | |
| Arginine | 0.10 | 0.19 | 0.045 | 0.16 | 0.44 (0.14-1.11) | 3.07 (0.19-28.29) | 0.11 | 0.43 |
| Glycine | 0.16 | 0.18 | 0.030 | 0.039 | 1.42 (1.14-1.79) | 1.96 (1.36-2.77) | 0.0024 | $2.1 \times 10^{-4}$ |
| Histidine | 0.32 | 0.23 | 0.34 | 0.19 | 2.09 (1.69-2.60) | 1.55 (1.37-1.76) | $1.8 \times 10^{-11}$ | $1.0 \times 10^{-11}$ |
| Phenylalanine | 0.27 | 0.21 | 0.12 | 0.14 | (reference) | | | |
| Serine | 0.15 | 0.19 | 0.39 | 0.33 | 2.33 (1.89-2.91) | 1.06 (0.76-1.46) | $2.0 \times 10^{-14}$ | 0.73 |
| Tyrosine | 0.0024 | 0.0030 | 0.079 | 0.15 | 1.24 (0.37-3.03) | 0.60 (0.47-0.77) | 0.68 | $5.8 \times 10^{-5}$ |
| HLA-DRB1 amino acid position 71 | | | | | | | | |
| Alanine | 0.10 | 0.18 | 0.04 | 0.15 | 1.96 (0.75-6.43) | 0.24 (0.026-3.98) | 0.21 | 0.32 |
| Arginine | 0.82 | 0.73 | 0.33 | 0.45 | (reference) | | | |
| Glutamic acid | 0.073 | 0.074 | 0.083 | 0.12 | 2.37 (1.72-3.27) | 1.09 (0.84-1.39) | $1.1 \times 10^{-7}$ | 0.53 |
| Lysine | 0.0096 | 0.011 | 0.54 | 0.28 | 0.75 (0.43-1.19) | 1.85 (1.57-2.18) | 0.25 | $8.7 \times 10^{-14}$ |

HLA, human leucocyte antigen; OR, odds ratio; 95% CI, 95% confidence interval.
†Obtained from the multivariate regression model that included all the variants listed here.

**Supplementary Table 6. A correspondence table of amino acid polymorphisms and 4-digit classical HLA alleles**

A correspondence table of amino acid polymorphisms and 4-digit classical HLA alleles is provided in an excel file format. It includes 4-digit classical alleles with a frequency >1% in either Europeans or Japanese populations based on the reference panels.
